# Multi-organ impairment and Long COVID: a 1-year prospective, longitudinal cohort study

**DOI:** 10.1101/2022.03.18.22272607

**Authors:** Andrea Dennis, Daniel J Cuthbertson, Dan Wootton, Michael Crooks, Mark Gabbay, Nicole Eichert, Sofia Mouchti, Michele Pansini, Adriana Roca-Fernandez, Helena Thomaides-Brears, Matt Kelly, Matthew Robson, Lyth Hishmeh, Emily Attree, Melissa Heightman, Rajarshi Banerjee, Amitava Banerjee

## Abstract

**Importance:** Multi-organ impairment associated with Long COVID is a significant burden to individuals, populations and health systems, presenting challenges for diagnosis and care provision. Standardised assessment across multiple organs over time is lacking, particularly in non-hospitalised individuals.

**Objective:** To determine the prevalence of organ impairment in Long COVID patients at 6 and at 12 months after initial symptoms and to explore links to clinical presentation.

**Design:** This was a prospective, longitudinal study in individuals following recovery from acute COVID-19. We assessed symptoms, health status, and multi-organ tissue characterisation and function, using consensus definitions for single and multi-organ impairment. Physiological and biochemical investigations were performed at baseline on all individuals and those with organ impairment were reassessed, including multi-organ MRI, 6 months later.

**Setting:** Two non-acute settings (Oxford and London).

**Participants:** 536 individuals (mean 45 years, 73% female, 89% white, 32% healthcare workers, 13% acute COVID-19 hospitalisation) completed baseline assessment (median: 6 months post-COVID-19). 331 (62%) with organ impairment or incidental findings had follow up, with reduced symptom burden from baseline (median number of symptoms: 10 and 3, at 6 and 12 months).

**Exposure:** SARS-CoV-2 infection 6 months prior to first assessment.

**Main outcome:** Prevalence of single and multi-organ impairment at 6 and 12 months post-COVID-19.

**Results:** Extreme breathlessness (36% and 30%), cognitive dysfunction (50% and 38%) and poor health-related quality of life (EQ-5D-5L<0.7; 55% and 45%) were common at 6 and 12 months, and associated with female gender, younger age and single organ impairment. At baseline, there was fibro-inflammation in the heart (9%), pancreas (9%), kidney (15%) and liver (11%); increased volume in liver (7%), spleen (8%) and kidney (9%); decreased capacity in lungs (2%); and excessive fat deposition in the liver (25%) and pancreas (15%). Single and multi-organ impairment were present in 59% and 23% at baseline, persisting in 59% and 27% at follow-up.

**Conclusion and Relevance:** Organ impairment was present in 59% of individuals at 6 months post-COVID-19, persisting in 59% of those followed up at 1 year, with implications for symptoms, quality of life and longer-term health, signalling need for prevention and integrated care of Long COVID.

**Trial Registration:** ClinicalTrials.gov Identifier: NCT04369807

**Key points:**

- Question: What is the prevalence of organ impairment in Long COVID at 6- and 12-months post-COVID-19?
- Findings: In a prospective study of 536 mainly non-hospitalised individuals, symptom burden decreased, but single organ impairment persisted in 59% at 12 months post-COVID-19.
- Meaning: Organ impairment in Long COVID has implications for symptoms, quality of life and longer-term health, signalling need for prevention and integrated care of Long COVID.

## Introduction

Symptoms of Long COVID, also known as post-acute sequelae of COVID-19 (PASC), are well-documented^1,2^, but natural history is poorly characterised, whether by symptoms, organ impairment or function^3–7^. Among 3,762 individuals with suspected or confirmed COVID-19, fatigue, breathlessness and cognitive dysfunction were the most debilitating symptoms beyond 35 weeks^8^. In the UK’s largest Long COVID clinic, non-hospitalised patients required specialist referral at similar rates to hospitalised patients and were more likely to report breathlessness and fatigue, with reduced health-related quality of life (HRQoL)^8^. A US study in 270,000 individuals post-COVID-19 showed one-third had persistent symptoms at 3-6 months (more common than post-influenza symptoms), possibly due to direct organ-specific rather than general viral effects, and potentially informing development of effective treatments^9^.

Long COVID may be linked to severity of initial illness in some hospitalised patients, but prognostic factors are neither defined nor investigated systematically in non-hospitalised patients^8–10^. To conduct trials of possible therapies for Long COVID, we need stratification by symptoms or investigations^3^. Our preliminary MRI data in 201 individuals showed mild organ impairment in the heart, lungs, kidneys, liver, pancreas, and spleen, with single and multi-organ impairment in 70% and 29%, respectively, 4 months after COVID-19. Clinical utility of these MR metrics for chronic and multi-system conditions has been shown^11-12^. More severe ongoing symptoms of breathlessness and fatigue were associated with myocarditis (p<0.05)^10^, but symptoms and multi-organ manifestations have not been correlated.

In the UK and other countries, health system and research responses have begun at scale^13–15^. However, clinical patient pathways are unclear and there are still no proven, evidence-based therapies, whether in subgroups or in the overall Long COVID population. Single and multi-organ impairment need investigation over medium- and long-term to assess resource utilisation and health system needs.

In individuals with Long COVID, we therefore prospectively investigated:

1. Symptoms, organ impairment and function over 1 year, particularly relating to ongoing breathlessness, cognitive dysfunction and HRQoL
2. Associations between symptoms and organ impairment

## Methods

### Patient Population and Study Design

Recruitment was by response to advertisement or specialist referral to two non-acute sites (Perspectum, Oxford and Mayo Clinic Healthcare, London) from April 2020 to August 2021, based on prior SARS-CoV-2 infection and written informed consent. (Supplementary Methods). Exclusion criteria were active respiratory infection symptoms (temperature >37.8°C or ≥3 coughing episodes in 24 hours); hospital discharge in last 7 days; asymptomatic or hospital discharge ≥4 months prior to enrolment and contraindications to MRI (e.g. pacemakers, defibrillators, metallic implanted devices, claustrophobia). Those with evidence of organ impairment, based on bloods, MRI, or incidental findings, were invited for 6-month follow-up (contacted on ≥2 occasions to minimise loss to follow-up). Each visit comprised MRI and blood investigations (full blood count, biochemistry) and online questionnaires completed beforehand^10^.

### Diagnostic Assessment in Non-acute Settings

Quantitative multi-organ MRI (CoverScan, Perspectum, Oxford) was used to assess organ impairment as previously reported (Figure S1)^10^, using healthy controls (no prior COVID-19 diagnosis; 59 in Oxford, 33 in London). Participants underwent a 40-minute MRI scan of lungs, heart, kidney, liver, pancreas and spleen on 1.5T or 3T Siemens scanners at three imaging sites (Oxford: MAGNETOM Aera 1.5T, Mayo Healthcare London: MAGNETOM Vida 3T, Chenies Mews Imaging Centre London: MAGNETOM Prisma 3T). MR metrics were standardised to deliver a single report interpretable by clinicians. Each report included 49 organ-specific metrics with reference ranges to determine impairment (updated from our prior study^16^) after determining distribution of each metric in healthy controls matched for age and sex (n=92) and for organ volumes from healthy controls representing complete sex and height subgroups (N=1835) in this study and UK Biobank^17^ (Table S1a and b). Repeatability of the metrics was evaluated in healthy controls using standardized performance testing criteria. Technical success was determined by reporting quality-assured measures for each variable reported herein, and overall, in delivering a report for each patient.

### Symptoms, Function and Organ Impairment

Assessment focused on commonly reported symptoms, HRQoL and degree of breathlessness (Dyspnoea 12 scale^8^). HRQoL was assessed by validated EQ-5D-5L (EuroQOL), comprising: (1) five health dimensions (mobility, self-care, usual activities, pain and discomfort, and anxiety and depression) each at five severity levels (none, slight, moderate, severe, and extreme); (2) self-rated health using visual analogue scale (VAS) from 0 (worst imaginable) to 100 (best imaginable); (3) derived EQ-5D ‘utility’ or index score from 0 (‘dead’) to 1 (‘full health’)^10^. Participants were asked at follow-up about time off work due to COVID-19 (not done at baseline). Multi-organ impairment at baseline and follow-up was defined as ≥2 MRI measurements from different organs outside reference ranges. Elevations in liver or kidney volume were excluded from definitions of organ impairment^10^. Further details are in Supplementary Methods.

### Associations Between Symptoms and Organ Impairment

The most common symptoms were classified according to published subgroups^8^: systemic (fever, myalgia, joint pain, fatigue/malaise, headaches), cardiopulmonary (wheezing, chest pain, shortness of breath), cognitive dysfunction (poor memory, cognition and concentration), poor HRQoL (EQ-5D utility score<0.7) and a subset of the cardiopulmonary subgroup for severe breathlessness (dyspnoea-12 total score≥10). Fisher’s exact or Kruskal Wallis tests were used for categoric and continuous data, comparing: (i) each of the 5 symptom groups vs. those with none of these symptoms, (ii) asymptomatic vs symptomatic at follow-up, and (iii) baseline vs follow-up or participants vs controls. Stepwise multi-variable logistic regression was performed for associations with 5 symptom groups at baseline or follow-up. Since pre-COVID data were unavailable, organ impairment may pre-date COVID-19. A sensitivity analysis was conducted, excluding those at risk of metabolic disease unlikely to have manifested between first symptoms and baseline assessment (including BMI ≥30 kg/m^2^, hypertension and diabetes) (Supplementary methods).

### Statistical Analysis

Based on UK Biobank studies of organ impairment, required sample size for associations between COVID-19 and 10% prevalence of liver impairment was 507^18,19^. Analyses were in R version 4.1.0, defining statistical significance by uncorrected p-value<0.05(2-sided). Descriptive statistics were used for baseline characteristics; normally distributed-continuous variables: mean(standard deviation, SD); non-normally distributed-continuous: median(interquartile range, IQR), and categorical variables: frequency(percentage).

### Ethical Approval

The protocol was approved by a UK ethics committee (20/SC/0185) and registered (https://clinicaltrials.gov/ct2/show/NCT04369807).

## Results

### Study Population for Diagnostic Assessment in Non-Acute Settings

536 individuals (mean age 45 years, 73% female, median BMI 25 kg/m^2^, 13% COVID-19 hospitalisation, 32% healthcare workers) were included at baseline. 388 (72%) with organ impairment were eligible for follow-up, which 331 (62%) completed. Demographics were comparable to healthy controls (Table S1a). Most were ‘UK first wave’ (COVID-19 January-September 2020: n=497); 39 ‘second wave’ (COVID-19 after September 2020). Median time from initial COVID-19 symptoms to baseline assessment, and baseline to follow-up were 182 (IQR: 132-222) and 196 days (IQR: 182-209). Demographics and risk factors showed no major differences between baseline and follow-up (Figure 1, Table 1). Participants with persistent symptoms in the five symptom groups were more likely to be female and obese at baseline (Table 2, Table S2). Technical success of MRI and integrated in-person assessment was 99.1% and 98.3% at baseline and follow-up assessments. Reports were delivered to patients and primary care clinicians in all cases.

**Table 1:** Characteristics of patients at baseline and follow-up. P-values represent results from an independent unpaired t-test (or non-parametric equivalent), Fisher’s exact test for dichotomous variables and chi-square test for categories.

| Characteristic | Baseline<br>(n=536) | Follow-up<br>(n=331) | p-value |
| --- | --- | --- | --- |
| Age (years) | 45 (11) | 47 (11) | 0.003 |
| Sex (n females) | 389 (73%) | 241 (73%) | 1 |
| BMI (kg/m<sup>2</sup>) | 25 (23, 29) | 26 (23,31) | 0.03 |
| Ethnicity |  |  | 0.99 |
| White | 477 (89%) | 295 (89%) |  |
| Mixed | 6 (1%) | 3 (1%) |  |
| South Asian | 21 (4%) | 15 (5%) |  |
| Black | 10 (2%) | 5 (2%) |  |
| Healthcare worker | 172 (32%) | 112 (34%) | 0.603 |
| At least one COVID-19 vaccination | Not recorded | 197 (76%)<br>missing: n=71 |  |
| **Comorbidities and risks** |  |  |  |
| Smoking |  |  |  |
| Never | 349 (65%) | 222 (67%) | 0.606 |
| Current | 14 (3%) | 7 (2%) | 0.821 |
| Ex-smoker | 172 (32%) | 102 (31%) | 0.707 |
| BMI |  |  |  |
| ≥25 kg/m<sup>2</sup> | 293 (55%) | 200 (61%) | 0.090 |
| ≥30 kg/m<sup>2</sup> | 120 (22%) | 92 (28%) | 0.074 |
| Hypertension | 44 (8%) | 37 (11%) | 0.151 |
| Diabetes | 10 (2%) | 9 (3%) | 0.476 |
| Heart disease | 9 (2%) | 4 (1%) | 0.776 |
| Asthma | 101 (19%) | 56 (17%) | 0.525 |
| Hospitalised during acute COVID-19 | 72 (13%) | 57 (17%) | 0.141 |
| Time off work (days) | Not recorded | 125 (35, 296) | - |
| **15 common symptoms** |  |  |  |
| Number reported [median (IQR)] | 10 (8, 11) | 3 (0, 5) | <0.001 |
| None reported in history | 0 (0%) | 93 (28%) | <0.001 |
| None reported in history/ questionnaires | 0 (0%) | 60 (19%) | <0.001 |
| **Symptom groups** |  |  |  |
| Systemic | 245 (46%) | 2 (1%) | <0.001 |
| Cardiopulmonary | 238 (44%) | 15 (5%) | <0.001 |
| Severe breathlessness (Dyspnoea 12 ≥10) | 187 (36%) | 93 (30%) | 0.094 |
| Cognitive dysfunction | 268 (50%) | 127 (38%) | <0.001 |
| Poor HRQoL | 281 (55%) | 138 (45%) | 0.011 |
| None of five symptom groups | 66 (13%) | 108 (35%) | <0.001 |
| **Duration (days: median, [IQR])** |  |  |  |
| Initial symptoms-to-assessment | 182 (132, 222) | 384 (350, 431) | - |
| COVID-19 positive-to-assessment | 110 (53, 175) | 328 (265, 375) | - |

**Table 2:** Characteristics by symptom status at baseline and follow-up. Compared are the baseline characteristics (top part) and the follow-up characteristics (bottom part) between groups.

| Characteristic | Persistent symptoms (n=264) | Asymptomatic (n=60) | p-value |
| --- | --- | --- | --- |
| **Baseline characteristics** |  |  |  |
| **Demographics** |  |  |  |
| Age (years) | 45 (40, 53) | 47 (39, 58) | 0.33 |
| Female sex (n) | 204 (77%) | 34 (57%) | **0.002** |
| BMI (kg/m<sup>2</sup>) | 26 (23, 30) | 28 (23, 31) | 0.518 |
| White | 239 (91%) | 49 (82%) | 0.066 |
| Mixed | 3 (1%) | 0 (0%) | >0.999 |
| South Asian | 8 (3%) | 7 (12%) | **0.01** |
| Black | 3 (1%) | 2 (3%) | 0.232 |
| Healthcare worker | 89 (34%) | 19 (32%) | 0.88 |
| **Comorbidities and risks** |  |  |  |
| Never smoked | 173 (66%) | 40 (67%) | >0.999 |
| Current smoker | 4 (2%) | 2 (3%) | 0.308 |
| Ex-smoker | 87 (33%) | 18 (30%) | 0.76 |
| BMI ≥ 25 kg/m<sup>2</sup> | 159 (60%) | 38 (63%) | 0.77 |
| BMI ≥ 30 kg/m<sup>2</sup> | 72 (27%) | 18 (30%) | 0.75 |
| Hypertension | 25 (9%) | 6 (10%) | 0.813 |
| Diabetes | 3 (1%) | 4 (7%) | **0.024** |
| Asthma | 53 (20%) | 9 (15%) | 0.468 |
| Pre-existing heart disease | 3 (1%) | 1 (2%) | 0.561 |
| Hospitalized during acute COVID-19 | 46 (17%) | 10 (17%) | >0.999 |
| Number of common symptoms reported | 10 (9, 11) | 9 (7, 10) | **<0.001** |
| **MRI abnormality** |  |  |  |
| **Liver** |  |  |  |
| cT1 or fat high | 95 (36%) | 23 (39%) | 0.766 |
| cT1 (high) | 36 (14%) | 9 (15%) | 0.836 |
| cT1 (ms) | 727 (681, 770) | 715 (680, 768) | 0.689 |
| fat (high) | 87 (33%) | 20 (33%) | >0.999 |
| fat (%) | 3 (2, 6) | 3 (1, 6) | 0.907 |
| Volume (high) | 23 (9%) | 6 (10%) | 0.801 |
| Volume (ml) | 1,423 (1,252, 1,693) | 1,463 (1,277, 1,653) | 0.649 |
| **Pancreas** |  |  |  |
| sT1 or fat high | 64 (26%) | 14 (24%) | 0.868 |
| cT1 high | 29 (12%) | 7 (12%) | >0.999 |
| sT1 (ms) | 720 (686, 772) | 722 (684, 767) | 0.775 |
| Fat (high) | 51 (20%) | 12 (20%) | >0.999 |
| Fat (%) | 3 (2, 6) | 3 (3, 5) | 0.287 |
| **Kidney** |  |  |  |
| Cortex T1 (high) | 44 (17%) | 14 (24%) | 0.193 |
| Cortex T1 left 1.5T (ms) | 1,080 (1,041, 1,127) | 1,084 (1,046, 1,139) | 0.344 |
| Cortex T1 right 1.5T (ms) | 1,071 (1,028, 1,123) | 1,079 (1,042, 1,134) | 0.35 |
| Cortex T1 left 3T (ms) | 1,423 (1,380, 1,474) | 1,398 (1,360, 1,447) | 0.228 |
| Cortex T1 right 3T (ms) | 1,380 (1,350, 1,457) | 1,391 (1,311, 1,433) | 0.542 |
| Volume (high) | 21 (8%) | 7 (12%) | 0.318 |
| Volume left (ms) | 146 (129, 165) | 158 (133, 179) | **0.044** |
| Volume right (ms) | 146 (128, 166) | 150 (137, 172) | 0.184 |
| **Spleen** |  |  |  |
| Splenomegaly | 28 (11%) | 5 (8%) | 0.813 |
| Volume (ml) | 180 (141, 242) | 194 (166, 252) | 0.146 |
| **Lung** |  |  |  |
| FAC (low) | 7 (3%) | 0 (0%) | 0.359 |
| FAC (%) | 43 (10) | 42 (10) | 0.814 |
| **Heart** |  |  |  |
| Myocardial injury | 53 (20%) | 15 (25%) | 0.484 |
| T1 (high) | 24 (9%) | 11 (18%) | 0.062 |
| T1 1.5T (ms) | 980 (965, 993) | 972 (949, 987) | **0.047** |
| T1 3T (ms) | 1,185 (1,166, 1,199) | 1,184 (1,171, 1,214) | 0.922 |
| LV EF (low) | 13 (5%) | 4 (7%) | 0.531 |
| LV EF (%) | 60 (57, 63) | 58 (54, 61) | **0.025** |
| RV EF (low) | 10 (4%) | 4 (7%) | 0.302 |
| RV EF (%) | 59 (5) | 58 (6) | 0.108 |
| LV EDV (high) | 2 (1%) | 0 (0%) | >0.999 |
| LV EDF (ml) | 78 (71, 87) | 82 (75, 94) | **0.009** |
| RV EDV (high) | 3 (1%) | 0 (0%) | >0.999 |
| RV EDF (ml) | 74 (66, 84) | 80 (70, 93) | **0.005** |
| Global longitudinal strain (high) | 11 (4%) | 0 (0%) | 0.133 |
| Global longitudinal strain (%) | -14 (2) | -14 (2) | 0.484 |
| **Number of organs impaired on MRI** |  |  |  |
| 0 | 84 (32%) | 17 (28%) | 0.646 |
| ≥1 | 180 (68%) | 43 (72%) | 0.542 |
| ≥2 | 75 (28%) | 20 (33%) | 0.529 |
| ≥3 | 26 (10%) | 7 (12%) | 0.632 |
| **Follow-up characteristics** |  |  |  |
| Hospitalized between baseline and follow-up | 13 (5%) | 1 (2%) | 0.48 |
| Time off work (days) | 172 (42, 300) | 54 (21, 120) | **<0.001** |
| **MRI abnormality** |  |  |  |
| **Liver** |  |  |  |
| cT1 or fat high | 88 (35%) | 17 (28%) | 0.447 |
| cT1 (high) | 40 (16%) | 7 (12%) | 0.547 |
| cT1 (ms) | 724 (693, 768) | 708 (676, 755) | 0.15 |
| fat (high) | 74 (29%) | 16 (27%) | 0.754 |
| fat (%) | 3 (2, 6) | 3 (2, 6) | 0.876 |
| Volume (high) | 24 (9%) | 5 (8%) | >0.999 |
| Volume (ml) | 1,427 (1,271, 1,699) | 1,474 (1,316, 1,675) | 0.429 |
| **Pancreas** |  |  |  |
| sT1 or fat high | 43 (22%) | 10 (21%) | >0.999 |
| cT1 high | 16 (8%) | 4 (8%) | >0.999 |
| sT1 (ms) | 720 (684, 754) | 700 (673, 740) | 0.113 |
| Fat (high) | 35 (18%) | 10 (20%) | 0.683 |
| Fat (%) | 3 (2, 5) | 3 (2, 6) | 0.988 |
| **Kidney** |  |  |  |
| Cortex T1 (high) | 45 (17%) | 9 (15%) | 0.848 |
| Cortex T1 left 1.5T (ms) | 1,093 (1,047, 1,122) | 1,072 (1,047, 1,132) | 0.853 |
| Cortex T1 right 1.5T (ms) | 1,081 (1,048, 1,114) | 1,074 (1,033, 1,130) | 0.695 |
| Cortex T1 left 3T (ms) | 1,412 (1,370, 1,486) | 1,453 (1,402, 1,461) | 0.913 |
| Cortex T1 right 3T (ms) | 1,406 (1,356, 1,456) | 1,398 (1,356, 1,418) | 0.602 |
| Volume (high) | 21 (8%) | 6 (10%) | 0.61 |
| Volume left (ms) | 145 (127, 165) | 158 (137, 180) | **0.011** |
| Volume right (ms) | 149 (129, 167) | 156 (143, 172) | **0.028** |
| **Spleen** |  |  |  |
| Splenomegaly | 24 (9%) | 5 (8%) | >0.999 |
| Volume (ml) | 176 (139, 246) | 192 (161, 245) | 0.074 |
| **Lung** |  |  |  |
| FAC (low) | 4 (2%) | 0 (0%) | 0.58 |
| FAC (%) | 44 (11) | 45 (10) | 0.537 |
| **Heart** |  |  |  |
| Myocardial injury | 52 (20%) | 16 (28%) | 0.214 |
| T1 (high) | 25 (9%) | 7 (12%) | 0.632 |
| T1 1.5T (ms) | 982 (969, 992) | 975 (960, 987) | 0.069 |
| T1 3T (ms) | 1,181 (1,165, 1,206) | 1,185 (1,172, 1,200) | 0.974 |
| LV EF (low) | 6 (2%) | 1 (2%) | >0.999 |
| LV EF (%) | 60 (4) | 59 (5) | **0.03** |
| RV EF (low) | 7 (3%) | 5 (8%) | 0.05 |
| RV EF (%) | 59 (5) | 57 (5) | **0.018** |
| LV EDV (high) | 2 (1%) | 0 (0%) | >0.999 |
| LV EDF (ml) | 78 (70, 86) | 82 (75, 93) | **0.013** |
| RV EDV (high) | 4 (2%) | 0 (0%) | >0.999 |
| RV EDF (ml) | 74 (67, 86) | 81 (71, 91) | **0.017** |
| Global longitudinal strain (high) | 15 (6%) | 5 (9%) | 0.389 |
| Global longitudinal strain (%) | -15 (2) | -14 (2) | **0.022** |
| **Number of organs impaired on MRI** |  |  |  |
| 0 | 112 (42%) | 23 (38%) | 0.664 |
| ≥1 | 152 (58%) | 37 (62%) | 0.664 |
| ≥2 | 73 (28%) | 13 (22%) | 0.419 |
| ≥3 | 28 (11%) | 5 (8%) | 0.813 |

**Figure 1:**
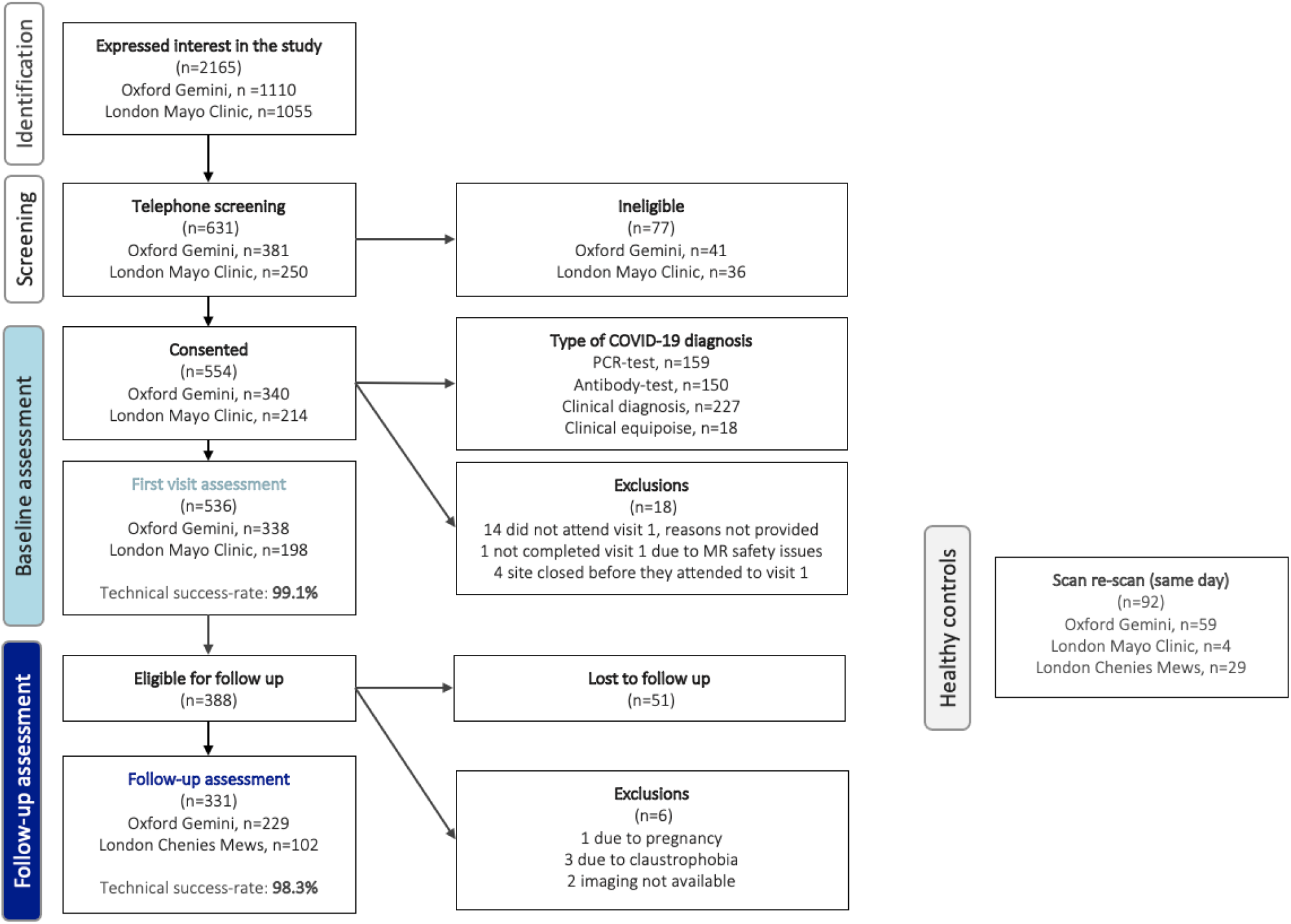
Study population from recruitment to follow-up.

### Symptoms over 1 Year

At baseline and follow-up, participants reported a median 10 (IQR: 8-11) symptoms and 3 (0-5) symptoms (p<0.001), respectively. At baseline, all five symptom groups had similar prevalence (systemic: 46%, cardiopulmonary: 44%, severe breathlessness: 36%, cognitive dysfunction: 50%, poor HRQoL: 55%); 13% reported none of these symptoms. At follow-up, symptoms were reduced, particularly systemic (1%) and cardiopulmonary (5%) (p<0.001). Exceptions were fatigue, breathlessness and cognitive dysfunction, where prevalence remained high. Some participants reported these symptoms only at follow-up. Females and individuals with obesity were more likely to have ≥1 of systemic symptoms, cardiopulmonary symptoms or poor HRQoL (p<0.001, 0.006 and <0.001 for sex and p=0.002, 0.012 and 0.004 for BMI). Most common symptoms improved by follow-up: fatigue (98% to 64%), myalgia (89% to 35%), shortness of breath (89% to 47%), headache (84% to 34%), chest pain (82% to 38%), fever (71% to 2%), cough (75% to 11%) and sore throat (72% to 11%). 60/331(18%) were asymptomatic at follow-up (Tables 1 and S2, Figure 2A).

**Figure 2:**
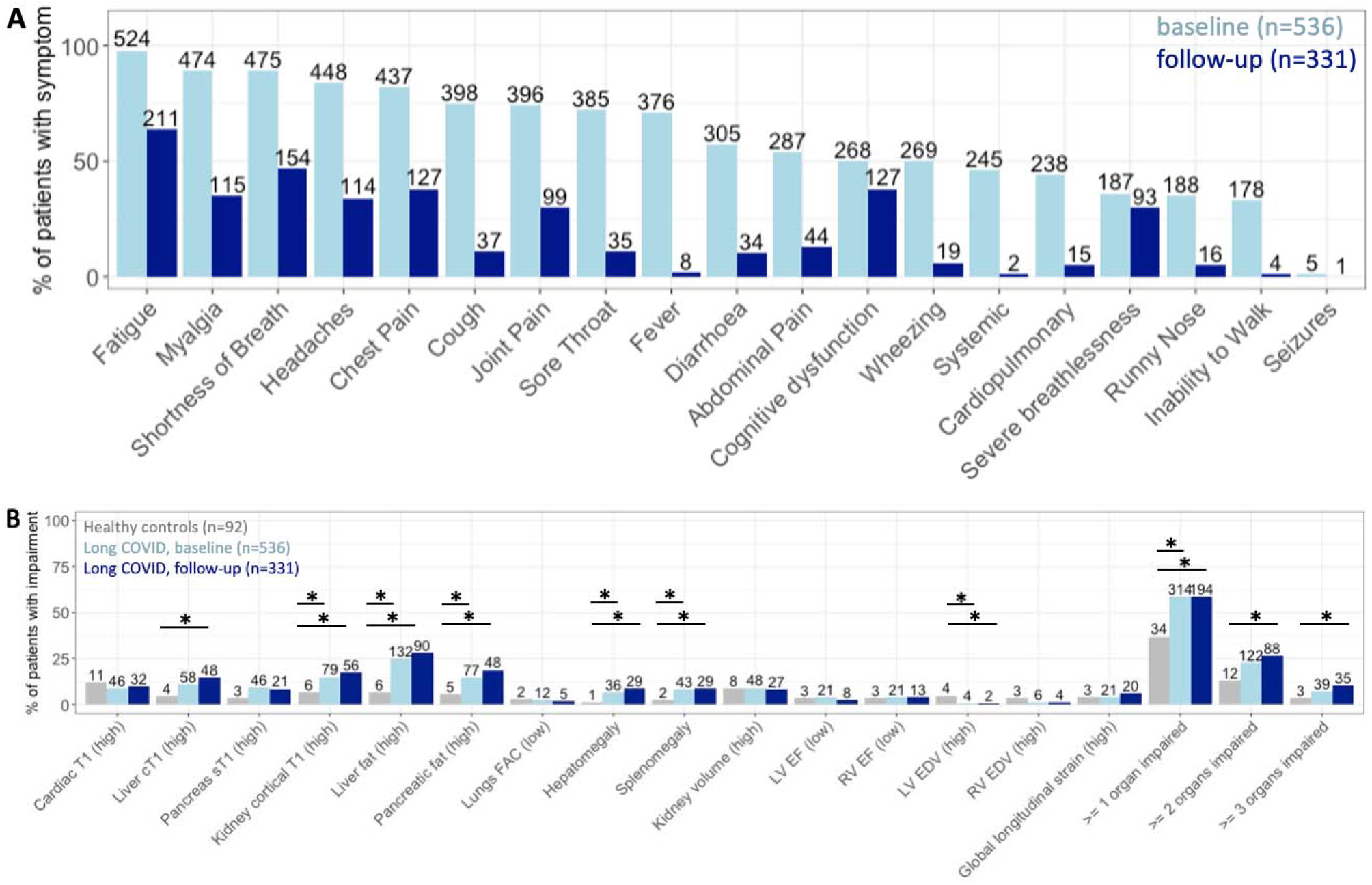
Proportion of individuals with Long COVID at baseline and follow-up for: (A) symptoms; (B) organ impairment. Note for Figure 2B: compared between healthy controls (grey) and at baseline and follow-up (blue). Significant differences (p<0.05) are indicated with a star, numbers above columns indicate the sample size (n) for each group.

### Function over 1 Year

HRQoL was poor at baseline: median VAS score 60% (40-70%) and median reported health utility index score 0.67 (IQR: 0.48-0.77) (Figure S2). The most highly ranked sub-optimal health dimensions were problems completing usual activities of living and pain (58% and 47% reporting moderate to extreme difficulties) (Figure S3). At follow-up, there was increased health utility index score to 0.71 (0.56, 0.81) (p<0.01), but 45% still reported utility score <0.7. At 12 months, 30% still complained of severe breathlessness. Almost every participant (281/302, 93%) took COVID-19-related time off work (median: 150 days IQR [45-300]). 97% of healthcare workers took time off work (median 180 days [41-308]) and 62 (63%) took >100 days off work. Those with ongoing symptoms reported taking more time off work (median 172 days [42-300]) compared to those who recovered (median 54 days [42-300]) (Table 2).

### Organ Impairment over 1 Year

Most standard-of-care biochemical investigations were within normal range and not predictive of outcomes (Table S3), except lactate dehydrogenase (80/500 [16%] and 70/319 [22%]), creatinine kinase (40/508 [8%] and 41/323 [13%], p=0.03); cholesterol (236/508 [46%] and 157/326 [48%]); and mean cell haemoglobin concentration (106/508 [21%] and 49/324 [15%]) which were elevated at baseline and follow-up.

On MRI of 536 participants at baseline, 59% and 23% had impairment in ≥1 and ≥2 organs respectively (Figure 2b), although impairment was usually mild (Table S4), e.g., among participants with cardiac impairment, none had severe heart failure. Organ impairment between visits did not improve significantly, unlike symptoms (Figure 2a). Participants without organ impairment had lowest symptom burden. At baseline, lung impairment (lower fractional area change) and impairment in ≥3 organs were associated with high symptom burden. At 12 months, there was reduction in symptom burden, except fatigue, breathlessness and reduced HRQoL where prevalence was unchanged (Figure S4). Healthcare workers were more likely to have liver impairment (p=0.017 at baseline and p=0.044 at follow-up) than the rest of the cohort.

In participants without symptoms at follow-up (n=60), organ impairment was present in 43 (72%) at baseline and 37 (62%) at follow-up (Table 2). In symptomatic participants at follow-up (n=264), there were 84 (32%) and 112 (42%) without organ impairment at baseline and follow-up, respectively. Liver steatosis, kidney fibro-inflammation and splenomegaly at baseline were more frequent in all symptom groups. Liver steatosis was associated with systemic symptoms and severe breathlessness (Table S2, Figure S5).

### Associations Between Symptoms and Organ Impairment

Looking at individual symptoms and by five symptom groups, neither abnormal biochemical investigations nor organ impairment were predictive of full recovery at follow-up (Tables S3-S6). Participants with normal MRI assessment had fewer baseline symptoms (Figure 3). Several liver-specific parameters were associated with specific symptom burden. High liver fat was present in 58/187 with severe breathlessness but only 70/257 without severe breathlessness at baseline. Conversely, low liver fat was more likely in those without severe breathlessness at both timepoints. High liver volume at follow-up was associated with lower HRQoL. Hepatomegaly was present in 20/138 with poor QoL but in only 8/167 with better QoL at follow-up. High liver fibro-inflammation was associated with cognitive dysfunction at follow-up: in 19% (24/124) with cognitive dysfunction but in only 12% (24/199) without cognitive dysfunction at follow-up (Table S6).

**Figure 3:**
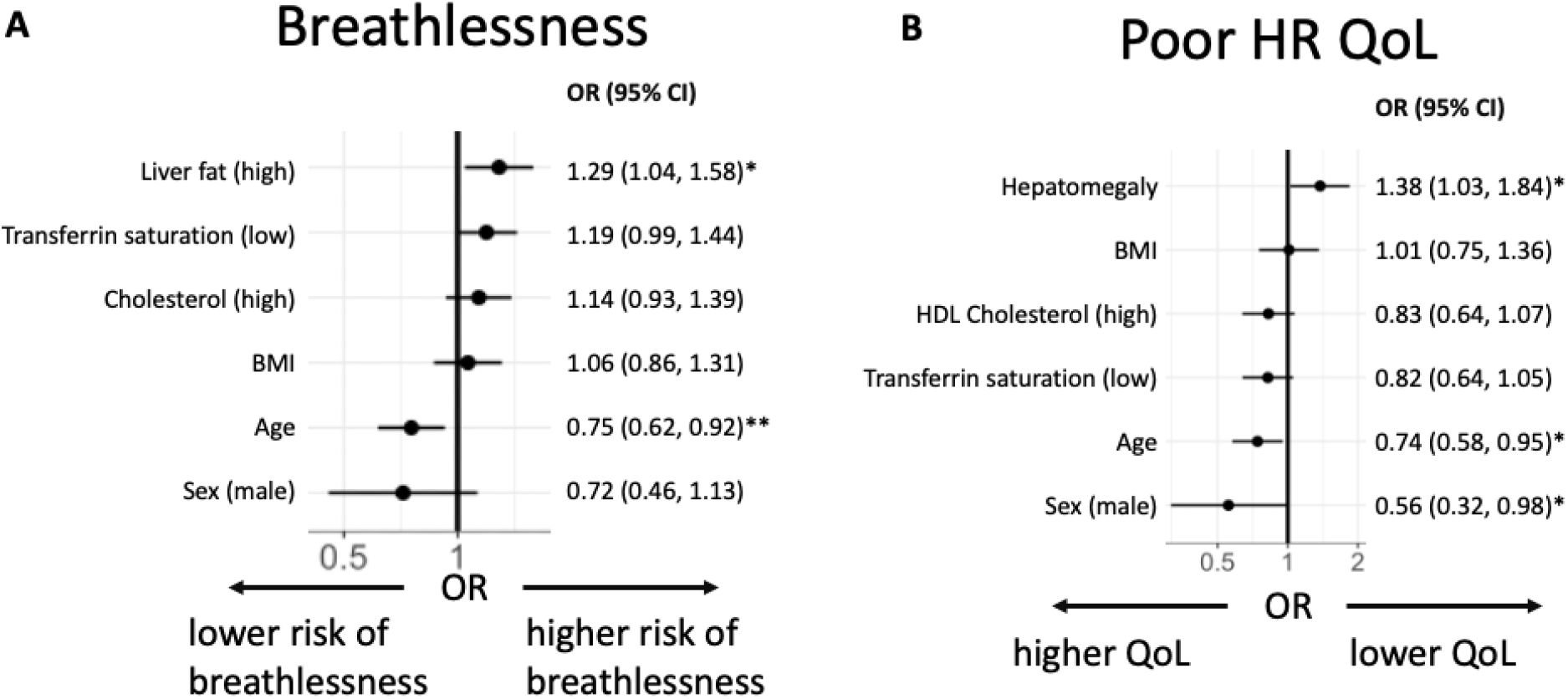
Association between risk factors and: (A) severe breathlessness at baseline; (B) Poor HRQoL at follow-up.

## Discussion

In this UK prospective study of largely non-hospitalised Long COVID, we have four new findings. First, we confirm multi-organ impairment at 6- and 12-months in 59% of individuals with Long COVID, with persistent symptoms and reduced function. Second, despite some associations between organ impairment and symptoms, there is currently insufficient evidence for distinct Long COVID subtypes. Blood biomarkers, the current standard of care, showed no relation to clinical presentation. Third, symptoms, blood investigations and quantitative, multi-organ MRI did not predict trajectory or recovery. Fourth, we demonstrate feasibility of scalable, multi-organ assessment in non-acute settings in the pandemic context.

Several studies confirm persistence of symptoms in individuals with Long COVID up to 1 year^20–22^. We now add that 3 in 5 people with Long COVID have impairment in at least one organ, and 1 in 4 have impairment in two or more organs, in some cases without symptoms. Impact on quality of life and time off work, particularly in healthcare workers, is a major concern for individuals, health systems and economies^23^. Many healthcare workers had no prior illness (2% diabetes, 2% heart disease and 22% asthma, which may play a pathophysiologic role^24^), but of 172 participants, 19 were still symptomatic at follow-up and off work at a median of 180 days. We need comparison with similar analyses from other Long COVID studies, and other long-term conditions^25^, with considerable workforce planning implications. There have been heterogenous methods to investigate Long COVID, whether qualitative^26–30^ or quantitative^7^, or symptom surveys^31^ versus cohort studies^4,32^. Most research still focuses on individual organs^33–36^. The scale of Long COVID burden necessitates action to develop, evaluate and implement evidence-based investigation, treatment and rehabilitation^37^ (e.g., STIMULATE-ICP and other studies^4^).

Metabolic disease, including non-alcoholic liver disease and diabetes, are postulated to play a role in pathogenesis of severe COVID-19 and possibly Long COVID^38^. We observe associations with markers of liver dysfunction, and increased risk of symptoms in female and obese individuals with Long COVID, which has been shown previously^39–41^. Cognitive impairment appears to develop in the natural history of Long COVID in some patients. As it is also associated with liver inflammation, it may be that, as with hepatic encephalopathy, disordered liver metabolism and gluconeogenesis may cause abnormal brain function. However, underlying mechanisms of the condition or syndrome remain elusive. We did not find evidence by symptoms, blood investigations or MRI to clearly define Long COVID subtypes.

We observed symptom recovery at 12 months in 30% of individuals with Long COVID, particularly with cardiopulmonary and systemic symptoms, aligning with other longitudinal studies^7,18,26^. Our findings of reductions in HRQoL and time at work reinforce prior research and are concerning, despite improvement over time. Future research must consider associations between symptoms, multi-organ impairment and function in larger cohorts, enabling clearer stratification and evaluation of treatments.

The COVERSCAN study was initiated in the early pandemic first wave when face-to-face assessment and investigation, and reduced health system capacity were major concerns for patients and health professionals. In the UK and other countries, Long COVID carries high burden of investigations and healthcare utilisation across specialties, and definitive care pathways are lacking. We show feasibility, acceptability and scalability of a rapid (40-minute), multi-organ MRI protocol for practice and research. Alongside routine clinical assessment and blood tests, COVERSCAN can exclude organ impairment in integrated, multidisciplinary care pathways^42^. Such MRI assessment has potential application beyond the pandemic for multi-system assessment and investigation, including in lower resource settings.

### Implications for Research

There are three research implications. First, complex intervention trials, including care pathways from investigation to rehabilitation are needed to evaluate therapies. Second, associations between symptoms, symptom groups, blood investigations and MRI must be investigated in larger populations. Third, Long COVID pathophysiology is still unclear, and should ideally be studied at the same time as clinical trials.

### Implications for Clinical Practice and Public Health

There are three practice and policy implications. First, CoverScan could be used to rule out organ impairment and to identify subgroups requiring specialist referral. Second, Long COVID is a multi-organ condition, needing multi-organ assessment and multi-disciplinary care. Third, poor 1-year post-COVID recovery rates highlight need for rehabilitation and integrated care, relevant to other long-term conditions.

### Strengths and Limitations

To date, this is the largest, comprehensive, systematic, multi-organ, post-COVID study over 1 year. The study cohort is representative of UK Long COVID populations in terms of risk factors and acute COVID-19 hospitalisation^8,9^. There are limitations. We did not have history and imaging prior to the pandemic and so it is difficult to determine if COVID-19/Long Covid caused impairment. There was no assessment of brain function. Not all participants had a laboratory-confirmed COVID-19 diagnosis. 38% of participants were not followed up. Generalizability of results from the UK’s first COVID-wave to the global population requires further exploration. We successfully delivered multi-organ MRI to assess organ health, but clinical utility remains to be determined. Health utilisation (e.g. primary care interactions) and economic burden of Long COVID were not assessed. Time off work was not assessed at baseline when the UK was in lockdown with a national furlough scheme in operation. Patients with normal assessment at baseline were not followed up due to resource and funding constraints. Therefore, whilst 222/536 (41%) patients at baseline were defined as normal after assessment, we cannot say they had better outcomes. Nevertheless, we can model cost-saving of a single assessment versus multiple assessments to achieve a discharge decision.

## Conclusions

Long COVID symptoms commonly persist at 12 months, even in those not severely affected by acute COVID-19, predominantly observed in women. Diagnosis and follow up of Long COVID can be performed in non-acute settings. Continued research in multi-system assessment and pharmacotherapy for those reporting ongoing fatigue, breathlessness, and cognitive problems is required to address Long COVID burden, in parallel with mechanistic studies to understand pathophysiology.

## Data Availability

Data are available upon reasonable request from the corresponding author.

## Acknowledgements

The study was partly financed by an Amendment to a European Commission’s Horizon 2020 grant 719445 (AMENDMENT Reference No AMD-719445-8): Non-invasive rapid assessment of chronic liver disease using Magnetic Resonance Imaging with LiverMultiScan (RADIcAL). AB has received funding from NIHR (including the STIMULATE-ICP study), AstraZeneca, European Union, UK Research and Innovation and British Medical Association. DW is funded by an NIHR advanced fellowship. MG is funded by ARC and DH&SC NIHR.

