## Supplementary methods, tables and figures for "Multi-organ impairment and Long COVID: a 1-year prospective, longitudinal cohort study"

| **Page Number** | **Contents** |
| --- | --- |
| 2 | Supplementary Methods |
| 5 | Table S1a: Demographics at baseline for those recovering from COVID-19 vs the controls used to derive reference ranges (median [IQR] or count [%]). |
| 6 | Table S1b: Reference ranges for MRI metrics. |
| 8 | Table S2: Characteristics for individuals with Long COVID with five major symptom categories at baseline, compared to those without any of those symptoms. |
| 10 | Table S3: Blood investigations at baseline and follow-up in individuals with Long Covid. |
| 12 | Table S4. Blood investigations by symptom status at baseline and follow-up in individuals with Long Covid. |
| 14 | Table S5: Comparison of MRI metrics between healthy controls and long COVID cohort at baseline. |
| 15 | Table S6: Associations between biomarkers and symptom groups in individuals with Long COVID. |
| 16 | Figure S1: Example MRI data segmentations used in organ morphology measurements. |
| 17 | Figure S2: Self-reported health related quality of life as reported from the EQ-5D-5L instrument showing the UK specific index scores (left) and the visual analogue score (right). |
| 18 | Figure S3: Dimensions of health from the EQ-5D-5L questionnaire. |
| 19 | Figure S4: Heat maps showing the proportion of those with impairment in individual organs that reported specific symptoms at baseline (left) and follow-up (right). |
| 20 | Figure S5: Proportion of cases with liver steatosis by symptom group (systemic, cardiopulmonary, severe breathlessness, brain fog, poor HRQoL). |
| 21 | Supplementary references |

**Supplementary Methods**

**SARS-CoV-2 Diagnosis**

Prior SARS-CoV-2 infection was defined based on a laboratory-confirmed tests (positive oro-/nasopharyngeal SARS-CoV-2 swab test by reverse-transcriptase-polymerase-chain reaction, or antibody test). Furthermore, strong clinical suspicion of infection (typical symptoms and COVID-19 diagnosis by two independent clinicians) was included as criterion, as testing was not widely available to non-hospitalized patients at the time of study recruitment. In n=18 patients, clinical diagnosis was performed only by one clinician (clinical equipoise).

**Imaging Acquisitions**

•	Cardiac MR imaging involved complete coverage of the heart with a short-axis stack (from the apex to the valve plane) of cine images acquired using cardiac gating, each acquired within a short breath-hold. This acquisition mirrors the one used at the UK Biobank and is a standardized approach [1]. Three short-axis and two long axis cine (HLA and VLA) cardiac T1 maps were also acquired using the MOLLI-T1 approach at the basal, mid, and apical levels of the left ventricle. Three short-axis and two long axis cine (HLA and VLA) at the basal, mid, and apical levels. For cardiac T2 maps, acquired using the fast low angle shot (FLASH) in 3T, at 1.5T scanner, TrueFISP was used (a Siemens version of balanced SSFP).

•	Liver and pancreas imaging used the LiverMultiScan acquisition protocol (Perspectum, Oxford, UK), which involves 3 single 2D axial slice breath-held acquisitions that separately are sensitive to the fat content (proton density fat fraction [PDFF]), to T2* (which can yield liver iron content) and a MOLLI-T1 measurement (providing a measurement of tissue water).Additionally, a volumetric scan was used that covers the entire liver [2].

•	Lungs: Two single slice dynamic cine MR acquisitions were acquired in the coronal plane with a 307ms temporal resolution: one 40 s acquisition with the patient instructed to breathe normally and a second 30 s acquisition with the patient instructed to breathe deeply.

•	Kidney: a single coronal view was used to image both kidneys. Imaging contrasts were MOLLI-T1, and a spoiled gradient recalled acquisition (spGR).

•	Spleen: Volumetric spGR MRI images

**Image Analysis**

•	Cardiac: Experienced cardiac MRI analysts used CVI42v5.11 (Cardiovascular Imaging Inc, Canada) to trace manually the myocardium in the end-diastolic and end-systolic phases in each of the short-axis views, following the standard UK Biobank evaluation approach as previously described [3]. We reported ventricular function; end systolic and diastolic volume; stroke volume and ejection fraction in both ventricles; left ventricular muscle mass and ventricular max wall thickness and global longitudinal and circumferential 3D strain metrics. Cardiac T1 and T2 measurements were determined as the mean of each of the 16 cardiac segments (of the AHA 17 segment model excluding the apex) [4]. When artefacts were present those measurements were not included in the results.

•	Liver Images were analysed by data analysts experienced at using the LiverMultiScan (Perspectum, Oxford, UK) software. This yielded global metrics in each liver of PDFF (proton density fat fraction), T2*, and cT1 (cT1 is a measurement of T1 that has been corrected for the confounding effects of iron and standardised to 3 Tesla; it is elevated with disease).

•	Pancreas images were analysed in an equivalent manner to the above except the software used was not FDA-cleared and iron correction was not performed [5]. The T1 was standardized to 3 Tesla.

•	Lung: Patient respiration was assessed by imaging a single 2D coronal slice of the lungs using a 30 second dynamic cine MRI acquisition (with a time resolution of 307ms), during which the patient was instructed to breathe deeply. In-house developed automated segmentation methods were used to segment the lungs and measure their areas in each time frame. Segmentations were reviewed and corrected by trained analysts. From these area vs time measurements, the lung area at maximum inspiration and expiration was measured, as well as a fractional area change (max area - min area)/max area.

•	Kidney: assessed using in-house tools to fit parametric maps and to allow trained analysts to make measurements. The kidney cortex was manually segmented using the MOLLI-T1 map to guide the boundary. Multiple regions-of-interests were manually placed within the cortex to extract a median value of cortical T1 in each kidney. Volumetric delineations of the kidneys were derived from SPGR MRI images. Automated delineations were produced using a 3D convolutional neural network, trained on expert annotations. Delineations were manually checked, and corrected, if necessary, for each subject.

•	Spleen: Volumetric delineations were derived from SPGR MRI images. Automated delineations were produced using a 3D convolutional neural network, trained on expert annotations. Delineations were manually checked, and corrected, if necessary, for each subject.

Technical failures were considered to be those acquisitions which did not produce images of sufficient quality as to derive metrics reported herein. For cardiac T1 and T2, this was based on derivation of global values for 3 segments.

**Reference Ranges**

All MRI metric reference ranges, but organ volumes, were calculated with n=92 Healthy Controls (HC) scanned at 1.5T and 3T based on the 2.5% (lower threshold) and 97.5% percentiles (upper threshold), using bootstrapping (100,000 permutations). Organ volumes were calculated from a combined cohort of 92 healthy controls and 1744 BMI-matched participants from the UK Biobank[6], representing all sex and height subgroups, as these are known confounders of organ size [7].

**Analysis**

**Organ Impairment**

Organ impairment was calculated for each organ based on evidence of any of the measurement appearing out of reference range (*Liver:* elevated cT1 or Fat; *Kidney:* elevated T1 or volume; *Pancreas:* elevated sT1 or Fat; *Heart:* elevated T1 in 3 or more segments, decreased RV or LV EF or increased LV or RV EDV or increased LV global longitudinal strain; *Spleen*: elevated volume; *Lung:* reduced fractional area volume). Thus, depending on the metric the upper or lower threshold was considered. For cardiac T1, elevation in at least 3 AHA segments defined elevated T1. Single organ impairment was based on at least 1 organ impaired and multi-organ impairment was based on 2 organ impairments.

**Associations of Symptoms and Biomarkers**

For multivariable regression, the outcomes of interest included separately the most common symptom groups systemic (reporting: fever, myalgia, joint pain, fatigue/malaise, headaches), cardiopulmonary (wheezing, chest pain, shortness of breath), severe breathlessness (dyspnoea-12 total score >=10), cognitive dysfunction (reporting any problems with memory, cognition and concentration), and poor HRQoL (EQ-5D utility score <0.7).

The regressors included i) demographic characteristics (age, sex, and BMI); ii) elevation in T1 metrics, steatosis or organ volume defined as a binary variable (liver cT1, PDFF and volume; pancreas sT1 and PDFF; kidney cortex T1 and volume, spleen volume, lung FAC, heart injury); iii) blood investigations outside of normal range in 15% of participants or significantly different between visits (cholesterol, HDL cholesterol, lactate dehydrogenase [LDH], mean corpuscular haemoglobin concentration [MCHC], potassium, low transferrin saturation, creatine kinase, basophils, low total iron binding capacity [TIBC], C-peptide) defined as binary variables. All continuous variables (BMI and age) were z-scored before entering the statistical models. Only observations with no missing data were included in the models.

For each outcome, first the effect of organ impairment was assessed in a multi-variable stepwise regression model, including regressors for demographics and all MRI organ metrics. Second, the effect of blood markers was assessed in a separate multi-variable stepwise regression model including demographic factors. Third, we performed a multivariable regression including regressors for demographic characteristics and those MRI or blood metrics found to be significant in the previous two models. Standardized odd ratios (OR) with 95% confidence intervals from this regression were presented in forest plots.

**Table S1a: Demographics at baseline for those recovering from COVID-19 vs the controls used to derive reference ranges (median [IQR] or count [%]).**

|  | **Long COVID** **n=536** | **control group 1,** **n=92** | **p-value*** | **control group 2,**  **n=1835** | **p-value**** |
| --- | --- | --- | --- | --- | --- |
| Sex (n female) | 389 (73%) | 61 (66%) | 0.3 | 1024 (56%) | **<0.001** |
| Age | 44 (38, 52) | 44 (32, 53) | 0.5 | 56 (49, 61) | **<0.001** |
| BMI | 26 (23, 29) | 23 (21, 25) | **<0.001** | 25 (23, 28)  (missing: n=3) | 0.2 |
| Ethnicity |  |  | 0.093 |  | **<0.001** |
| White | 477 (89%) | 85 (92%) |  | 1794 (98%) |  |
| Black | 13 (2%) | 0 (0%) |  | 5 (0%) |  |
| Chinese | 2 (0%) | 2 (2%) |  | 7 (0%) |  |
| South Asian | 22 (4%) | 16 (1%) |  | 13 (1%) |  |
| Other | 22 (4%) | 1 (1%) |  | 16 (1%) |  |
| Smoking status |  |  |  | (missing: n=3) | **0.021** |
| Current smoker | 14 (3%) | 3 (3%) | 0.7 | 101 (6%) |  |
| Past smoker | 172 (32%) | 17 (18%) | **0.012** | 591 (32%) |  |
| Hypertension | 44 (8%) | 0 (0%) | **0.009** | 0 (0%) | **<0.001** |
| Diabetic | 10 (2%) | 0 (0%) | 0.4 | 0 (0%) | **<0.001** |
| Heart disease | 9 (2%) | 2 (2%) | 0.08 | 65 (4%)  (missing: n=346) | **0.014** |
| Asthma | 101 (19%) | 0 (0%) | **<0.001** | 318 (23%)  (missing: n=435) | **<0.001** |

*p-value of comparisons between Long COVID patients and controls for reference ranges for all metrics but organ volumes (control group 1, n=92). **p-value of comparisons between Long COVID participants and a larger pool of healthy controls for organ volumes (control group 2, n=1836).

**Table S1b: Reference ranges for MRI metrics.**

|  | **Sex** | **Field Strength** | **Height (cm)** | **Lower threshold** | **Upper threshold (*)** |
| --- | --- | --- | --- | --- | --- |
| CARDIAC METRICS | | | | | |
| Field strength independent metrics (body-surface area [BSA] corrected) | | | | | |
| Left end diastolic volume (ml) | F | - | - | - | 108 |
| Left end diastolic volume (ml) | M | - | - | - | 132 |
| Right end diastolic volume (ml) | F | - | - | - | 110 |
| Right end diastolic volume (ml) | M | - | - | - | 139 |
| Field strength independent metrics (not BSA corrected) | | | | | |
| Global longitudinal strain 3D (%) | F | - | - | - | -11.45 |
| Global longitudinal strain 3D (%) | M | - | - | - | -7.75 |
| Left ventricle ejection fraction (%) | F | - | - | 52 | - |
| Left ventricle ejection fraction (%) | M | - | - | 51 | - |
| Right ventricle ejection fraction (%) | F | - | - | 50 | - |
| Right ventricle ejection fraction (%) | M | - | - | 50 | - |
| Field strength dependent metrics | | | | | |
| Global T1 ref range (ms) (#) | F | 1.5T | - | - | 1042 |
| Global T1 ref range (ms) (#) | M | 1.5T | - | - | 997 |
| Global T2 ref range (ms) (#) | - | 1.5T | - | - | 51 |
| Global T1 ref range (ms) (#) | F | 3T | - | - | 1255 |
| Global T1 ref range (ms) (#) | M | 3T | - | - | 1214 |
| Global T2 ref range (ms) (#) | - | 3T | - | - | 46 |
| LIVER METRICS | | | | | |
| Field strength independent metrics | | | | | |
| cT1 (ms) | - | - | - | - | 800 (*) |
| PDFF (%) | - | - | - | - | 5 (*) |
| Volume (ml) | F | - | < 164 | - | 1778 |
| Volume (ml) | M | - | < 164 | - | 2003 |
| Volume (ml) | F | - | ≥ 164  < 250 | - | 2048 |
| Volume (ml) | M | - | ≥ 164  < 250 | - | 2284 |
| KIDNEY METRICS | | | | | |
| Field strength independent metrics | | | | | |
| Left volume (ml) | F | - | < 164 | - | 177 |
| Left volume (ml) | M | - | < 164 | - | 221 |
| Left volume (ml) | F | - | ≥ 164  < 250 | - | 192 |
| Left volume (ml) | M | - | ≥ 164  < 250 | - | 255 |
| Right volume (ml) | F | - | < 164 | - | 176 |
| Right volume (ml) | M | - | < 164 | - | 207 |
| Right volume (ml) | F | - | ≥ 164  < 250 | - | 186 |
| Right volume (ml) | M | - | ≥ 164  < 250 | - | 229 |
| Field strength dependent metrics | | | | | |
| Left or Right Cortical T1 (ms) (§) | - | 1.5T | - | - | 1154 |
| Left or Right Cortical T1 (ms) (§) | - | 3T | - | - | 1512 |
| PANCREAS METRICS | | | | | |
| Field strength independent metrics | | | | | |
| sT1 (ms) | - | - | - | - | 821 |
| PDFF (%) | - | - | - | - | 6.6 (*) |
| SPLEEN METRICS | | | | | |
| Field strength independent metrics | | | | | |
| Volume (ml) | F | - | < 164 | - | 255 |
| Volume (ml) | M | - | < 164 | - | 392 |
| Volume (ml) | F | - | ≥ 164  < 250 | - | 293 |
| Volume (ml) | M | - | ≥ 164  < 250 | - | 411 |
| LUNG METRICS | | | | | |
| Field strength independent metrics | | | | | |
| Total deep fractional area change (%) | - | - | - | 22.0 | - |

(*) Reference ranges for the liver cT1 and liver PDFF were established from literature [8]. For pancreas PDFF, which has a positive skew in the distribution, reference ranges were extracted with the 95% percentile. (§) Right and left kidney cortex T1 limits were averaged for threshold setting. (#) Cardiac T1 and cardiac T2 were measured for each of 16 AHA segments but thresholds are reported for the global average.

**Table S2: Characteristics for individuals with Long COVID with five major symptom categories at baseline, compared to those without any of those symptoms.** P-value was for comparison between participants in a symptom group vs those with no symptoms.

|  | **No major symptoms** | **Systemic Symptoms** | | **Cardiopulmonary Symptoms** | | **Severe Breathlessness** | | **Cognitive Dysfunction** | | **Poor HRQoL** | |
| --- | --- | --- | --- | --- | --- | --- | --- | --- | --- | --- | --- |
|  | **n=66** | **n=245** | **p-value** | **n=238** | **p-value** | **n=187** | **p-value** | **n=268** | **p-value** | **n=281** | **p-value** |
| **Demographic characteristics** |  |  |  |  |  |  |  |  |  |  |  |
| Age | 45 (38, 51) | 44 (37, 52) | 0.948 | 43 (38, 51) | 0.557 | 43 (37, 50) | 0.285 | 44 (38, 52) | 0.792 | 43 (38, 51) | 0.613 |
| Female sex | 37/66 (56%) | 192/245 (78%) | **<0.001** | 178/238 (75%) | **0.006** | 143/187 (76%) | **0.003** | 201/268 (75%) | **0.004** | 224/281 (80%) | **<0.001** |
| BMI (kg/m^2^) | 25 (22, 27) | 26 (23, 31) | **0.007** | 26 (23, 30) | **0.015** | 26 (23, 30) | **0.017** | 25 (23, 30) | 0.072 | 25 (23, 30) | 0.082 |
| **Ethnicity** |  |  |  |  |  |  |  |  |  |  |  |
| White | 53/66 (80%) | 218/245 (89%) | 0.095 | 215/238 (90%) | **0.032** | 169/187 (90%) | **0.047** | 242/268 (90%) | **0.032** | 254/281 (90%) | **0.031** |
| Mixed | 0/66 (0%) | 3/245 (1.2%) | >0.999 | 3/238 (1.3%) | >0.999 | 4/187 (2.1%) | 0.575 | 3/268 (1.1%) | >0.999 | 5/281 (1.8%) | 0.588 |
| South Asian | 7/66 (11%) | 10/245 (4.1%) | 0.061 | 6/238 (2.5%) | **0.01** | 2/187 (1.1%) | **0.001** | 9/268 (3.4%) | **0.022** | 4/281 (1.4%) | **0.001** |
| Black | 3/66 (4.5%) | 3/245 (1.2%) | 0.112 | 3/238 (1.3%) | 0.119 | 3/187 (1.6%) | 0.185 | 4/268 (1.5%) | 0.142 | 5/281 (1.8%) | 0.18 |
| Health care worker | 17/66 (26%) | 87/245 (36%) | 0.145 | 71/238 (30%) | 0.544 | 56/187 (30%) | 0.636 | 87/268 (32%) | 0.373 | 87/281 (31%) | 0.457 |
| **Comorbidities and risks** |  |  |  |  |  |  |  |  |  |  |  |
| No smoker | 46/66 (70%) | 152/244 (62%) | 0.313 | 143/237 (60%) | 0.196 | 104/187 (56%) | 0.058 | 172/268 (64%) | 0.471 | 175/280 (62%) | 0.32 |
| Current smoker | 1/66 (1.5%) | 7/244 (2.9%) | >0.999 | 7/237 (3.0%) | >0.999 | 7/187 (3.7%) | 0.684 | 8/268 (3.0%) | >0.999 | 8/280 (2.9%) | >0.999 |
| Past smoker | 19/66 (29%) | 85/244 (35%) | 0.382 | 87/237 (37%) | 0.247 | 76/187 (41%) | 0.104 | 88/268 (33%) | 0.559 | 97/280 (35%) | 0.389 |
| BMI >25 kg/m^2^ | 34/66 (52%) | 141/245 (58%) | 0.404 | 136/238 (57%) | 0.484 | 104/187 (56%) | 0.569 | 141/268 (53%) | 0.891 | 148/281 (53%) | 0.892 |
| BMI >30 kg/m^2^ | 7/66 (11%) | 70/245 (29%) | **0.002** | 59/238 (25%) | **0.012** | 53/187 (28%) | **0.004** | 66/268 (25%) | **0.013** | 76/281 (27%) | **0.004** |
| Hypertension | 4/66 (6.1%) | 21/245 (8.6%) | 0.617 | 21/238 (8.8%) | 0.616 | 19/187 (10%) | 0.456 | 24/268 (9.0%) | 0.621 | 27/281 (9.6%) | 0.475 |
| Diabetes | 1/66 (1.5%) | 8/245 (3.3%) | 0.69 | 5/238 (2.1%) | >0.999 | 2/187 (1.1%) | >0.999 | 2/268 (0.7%) | 0.485 | 3/281 (1.1%) | 0.572 |
| Heart disease | 1/66 (1.5%) | 6/245 (2.4%) | >0.999 | 2/238 (0.8%) | 0.521 | 5/187 (2.7%) | >0.999 | 3/268 (1.1%) | 0.587 | 6/281 (2.1%) | >0.999 |
| Asthma | 8/66 (12%) | 45/245 (18%) | 0.272 | 59/238 (25%) | **0.029** | 45/187 (24%) | 0.052 | 52/268 (19%) | 0.211 | 57/281 (20%) | 0.16 |
| Hospitalized during acute COVID-19 | 11/66 (17%) | 41/245 (17%) | >0.999 | 37/238 (16%) | 0.849 | 20/187 (11%) | 0.274 | 31/268 (12%) | 0.299 | 37/281 (13%) | 0.434 |
| **Common symptoms** |  |  |  |  |  |  |  |  |  |  |  |
| Fever | 33/63 (52%) | 245/245 (100%) | **<0.001** | 179/238 (75%) | **<0.001** | 142/187 (76%) | **<0.001** | 189/268 (71%) | **0.007** | 210/281 (75%) | **<0.001** |
| Cough | 43/63 (68%) | 199/245 (81%) | **0.038** | 194/238 (82%) | **0.036** | 143/187 (76%) | 0.242 | 212/268 (79%) | 0.069 | 217/281 (77%) | 0.146 |
| Sore Throat | 37/63 (59%) | 191/245 (78%) | **0.003** | 190/238 (80%) | **<0.001** | 144/187 (77%) | **0.009** | 201/268 (75%) | **0.013** | 220/281 (78%) | **0.002** |
| Runny Nose | 21/63 (33%) | 84/245 (34%) | >0.999 | 97/238 (41%) | 0.312 | 74/187 (40%) | 0.453 | 102/268 (38%) | 0.563 | 101/281 (36%) | 0.771 |
| Wheezing | 10/63 (16%) | 137/245 (56%) | **<0.001** | 238/238 (100%) | **<0.001** | 126/187 (67%) | **<0.001** | 150/268 (56%) | **<0.001** | 156/281 (56%) | **<0.001** |
| Chest Pain | 34/63 (54%) | 214/245 (87%) | **<0.001** | 238/238 (100%) | **<0.001** | 172/187 (92%) | **<0.001** | 239/268 (89%) | **<0.001** | 251/281 (89%) | **<0.001** |
| Myalgia | 49/63 (78%) | 245/245 (100%) | **<0.001** | 216/238 (91%) | **0.008** | 171/187 (91%) | **0.007** | 242/268 (90%) | **0.01** | 256/281 (91%) | **0.007** |
| Joint Pain | 30/63 (48%) | 245/245 (100%) | **<0.001** | 189/238 (79%) | **<0.001** | 146/187 (78%) | **<0.001** | 207/268 (77%) | **<0.001** | 221/281 (79%) | **<0.001** |
| Fatigue | 59/63 (94%) | 245/245 (100%) | **0.002** | 237/238 (100%) | **0.007** | 184/187 (98%) | 0.07 | 267/268 (100%) | **0.005** | 276/281 (98%) | 0.062 |
| Shortness of Breath | 39/63 (62%) | 232/245 (95%) | **<0.001** | 238/238 (100%) | **<0.001** | 185/187 (99%) | **<0.001** | 250/268 (93%) | **<0.001** | 265/281 (94%) | **<0.001** |
| Inability to walk | 9/63 (14%) | 102/245 (42%) | **<0.001** | 95/238 (40%) | **<0.001** | 79/187 (42%) | **<0.001** | 90/268 (34%) | **0.002** | 110/281 (39%) | **<0.001** |
| Headaches | 37/63 (59%) | 245/245 (100%) | **<0.001** | 211/238 (89%) | **<0.001** | 160/187 (86%) | **<0.001** | 239/268 (89%) | **<0.001** | 252/281 (90%) | **<0.001** |
| Seizures | 0/63 (0%) | 4/245 (1.6%) | 0.585 | 1/238 (0.4%) | >0.999 | 2/187 (1.1%) | >0.999 | 3/268 (1.1%) | >0.999 | 4/281 (1.4%) | >0.999 |
| Abdominal pain | 21/63 (33%) | 157/245 (64%) | **<0.001** | 154/238 (65%) | **<0.001** | 121/187 (65%) | **<0.001** | 157/268 (59%) | **<0.001** | 180/281 (64%) | **<0.001** |
| Diarrhoea | 29/63 (46%) | 152/245 (62%) | **0.031** | 148/238 (62%) | **0.022** | 112/187 (60%) | 0.058 | 156/268 (58%) | 0.091 | 179/281 (64%) | **0.011** |
| Number of common symptoms | 7 (6, 9) | 11 (10, 12) | **<0.001** | 11 (10, 12) | **<0.001** | 11 (9, 12) | **<0.001** | 10 (9, 12) | **<0.001** | 11 (9, 12) | **<0.001** |
| **MRI abnormality** |  |  |  |  |  |  |  |  |  |  |  |
| Liver (cT1 or fat high) | 13/65 (20%) | 80/240 (33%) | **0.048** | 69/237 (29%) | 0.159 | 64/186 (34%) | **0.042** | 77/264 (29%) | 0.163 | 85/278 (31%) | 0.096 |
| Liver cT1 (high) | 8/64 (12%) | 30/238 (13%) | >0.999 | 25/236 (11%) | 0.655 | 21/185 (11%) | 0.822 | 30/263 (11%) | 0.828 | 36/277 (13%) | >0.999 |
| Liver fat (high) | 10/66 (15%) | 70/245 (29%) | **0.027** | 60/238 (25%) | 0.099 | 58/187 (31%) | **0.015** | 67/267 (25%) | 0.103 | 72/281 (26%) | 0.078 |
| Liver volume (high) | 5/66 (7.6%) | 19/244 (7.8%) | >0.999 | 16/237 (6.8%) | 0.787 | 19/187 (10%) | 0.632 | 18/267 (6.7%) | 0.788 | 24/280 (8.6%) | >0.999 |
| Pancreas (sT1 or fat high) | 12/63 (19%) | 51/230 (22%) | 0.729 | 43/224 (19%) | >0.999 | 41/177 (23%) | 0.597 | 47/252 (19%) | >0.999 | 53/264 (20%) | >0.999 |
| Pancreas cT1 (high) | 6/63 (9.5%) | 25/229 (11%) | >0.999 | 19/223 (8.5%) | 0.802 | 18/175 (10%) | >0.999 | 19/251 (7.6%) | 0.605 | 24/263 (9.1%) | >0.999 |
| Pancreatic fat (high) | 8/66 (12%) | 37/235 (16%) | 0.56 | 34/230 (15%) | 0.691 | 33/182 (18%) | 0.334 | 36/259 (14%) | 0.841 | 44/270 (16%) | 0.454 |
| Kidney (cortex T1 high) | 7/65 (11%) | 39/241 (16%) | 0.332 | 37/237 (16%) | 0.428 | 34/185 (18%) | 0.177 | 38/263 (14%) | 0.548 | 46/278 (17%) | 0.34 |
| Kidney volume (high) | 6/66 (9.1%) | 26/243 (11%) | 0.822 | 21/236 (8.9%) | >0.999 | 22/187 (12%) | 0.653 | 23/266 (8.6%) | >0.999 | 29/279 (10%) | >0.999 |
| Splenomegaly | 3/65 (4.6%) | 27/243 (11%) | 0.157 | 23/236 (9.7%) | 0.316 | 18/187 (9.6%) | 0.298 | 26/267 (9.7%) | 0.229 | 23/279 (8.2%) | 0.438 |
| Lung FAC (low) | 1/56 (1.8%) | 7/233 (3.0%) | >0.999 | 6/231 (2.6%) | >0.999 | 5/180 (2.8%) | >0.999 | 5/262 (1.9%) | >0.999 | 7/271 (2.6%) | >0.999 |
| Heart injury | 15/66 (23%) | 42/240 (18%) | 0.372 | 44/234 (19%) | 0.486 | 34/183 (19%) | 0.474 | 48/261 (18%) | 0.485 | 56/276 (20%) | 0.736 |
| Cardiac global T1 (elevated in >=3 segments) | 8/66 (12%) | 17/245 (6.9%) | 0.2 | 21/238 (8.8%) | 0.477 | 15/187 (8.0%) | 0.325 | 20/268 (7.5%) | 0.221 | 22/281 (7.8%) | 0.328 |
| LF EF (low) | 3/66 (4.5%) | 9/243 (3.7%) | 0.724 | 8/237 (3.4%) | 0.71 | 7/186 (3.8%) | 0.725 | 10/266 (3.8%) | 0.727 | 9/280 (3.2%) | 0.706 |
| RF EF (low) | 2/66 (3.0%) | 8/243 (3.3%) | >0.999 | 11/237 (4.6%) | 0.741 | 2/186 (1.1%) | 0.281 | 11/266 (4.1%) | >0.999 | 9/280 (3.2%) | >0.999 |
| LV EDV (high) | 1/66 (1.5%) | 1/243 (0.4%) | 0.382 | 2/237 (0.8%) | 0.523 | 1/186 (0.5%) | 0.456 | 2/266 (0.8%) | 0.487 | 1/280 (0.4%) | 0.346 |
| RV EDV (high) | 0/66 (0%) | 3/243 (1.2%) | >0.999 | 4/237 (1.7%) | 0.58 | 2/186 (1.1%) | >0.999 | 5/266 (1.9%) | 0.587 | 3/280 (1.1%) | >0.999 |
| Global longitudinal strain (high) | 3/66 (4.5%) | 10/240 (4.2%) | >0.999 | 7/233 (3.0%) | 0.464 | 8/183 (4.4%) | >0.999 | 9/258 (3.5%) | 0.715 | 17/273 (6.2%) | 0.775 |
| **Multi-organ** |  |  |  |  |  |  |  |  |  |  |  |
| no organ impaired | 29/66 (44%) | 95/245 (39%) | 0.48 | 94/238 (39%) | 0.571 | 66/187 (35%) | 0.238 | 114/268 (43%) | 0.89 | 113/281 (40%) | 0.581 |
| >= 1 organ impaired | 37/66 (56%) | 150/245 (61%) | 0.48 | 144/238 (61%) | 0.571 | 121/187 (65%) | 0.238 | 154/268 (57%) | 0.89 | 168/281 (60%) | 0.581 |
| >=2 organs impaired | 12/66 (18%) | 66/245 (27%) | 0.154 | 49/238 (21%) | 0.731 | 49/187 (26%) | 0.242 | 56/268 (21%) | 0.734 | 68/281 (24%) | 0.333 |
| >=3 organs impaired | 2/66 (3.0%) | 23/245 (9.4%) | 0.125 | 21/238 (8.8%) | 0.185 | 19/187 (10%) | 0.116 | 23/268 (8.6%) | 0.189 | 25/281 (8.9%) | 0.13 |

*.*

**Table S3: Blood investigations at baseline and follow-up in individuals with Long COVID.** The groups were compared using Fisher’s exact test.

|  | **Baseline**, n=536 | **Follow-up**, n=331 | **p-value*** | **Unit** | **Reference range** |
| --- | --- | --- | --- | --- | --- |
| Alanine transferase (high)  (low) | 74/508 (15%)  7/508 (1%) | 46/326 (14%)  4/326 (1%) | 0.92  >0.999 | IU/L | F: 10-35, M: 10-50 |
| Albumin (high)  (low) | 27/508 (5%)  0/508 (0%) | 11/326 (3%)  0/326 (0%) | 0.234 | g/L | 34-50 |
| Alkaline Phosphatase (high)  (low) | 13/508 (3%)  13/508 (3%) | 11/326 (3%)  5/326 (2%) | 0.528  0.465 | IU/L | F: 35-104, M: 40-129 |
| Amylase (high)  (low) | 34/463 (7%)  10/463 (2%) | 22/279 (8%)  10/279 (4%) | 0.776  0.251 | IU/L | 28 - 100 |
| Aspartate transferase (high)  (low) | 43/488 (9%)  0/488 (0%) | 37/312 (12%)  0/312 (0%) | 0.184 | IU/L | F: 0-31, M:0-37 |
| Basophils (high)  (low) | 2/508 (0%)  0/508 (0%) | 7/324 (2%)  0/324 (0%) | **0.032** | 10^9/L | 0.0-0.1 |
| Bicarbonate (high)  (low) | 25/508 (5%)  49/508 (10%) | 21/326 (6%)  33/326 (10%) | 0.355  0.813 | mmol/L | 22-29 |
| Bilirubin (high)  (low) | 17/508 (3%)  0/508 (0%) | 12/326 (4%)  0/326 (0%) | 0.847 | μmol/L | 0-20 |
| C-peptide (high)  (low) | 19/463 (4%)  0/463 (0%) | 24/279 (9%)  1/279 (0%) | **0.014**  0.376 | μg/L | 1.1 - 4.4 |
| Calcium (high)  (low) | 7/508 (1%)  8/508 (2%) | 1/326 (0%)  5/326 (2%) | 0.159  >0.999 | mmol/L | 2.20-2.60 |
| Chloride (high)  (low) | 10/508 (2%)  11/508 (2%) | 1/326 (0%)  6/326 (2%) | 0.058  0.807 | mmol/L | 98-107 |
| Cholesterol (high) | 236/508 (46%) | 157/326 (48%) | 0.67 | mmol/L | <5 |
| Creatine kinase (high)  (low) | 40/508 (8%)  2/508 (0%) | 41/323 (13%)  2/323 (1%) | **0.03**  0.644 | IU/L | F: 26-104, M: 38-204 |
| Creatinine (high)  (low) | 6/508 (1%)  25/508 (5%) | 6/326 (2%)  16/326 (5%) | 0.553  >0.999 | μmol/L | F: 49-92, M: 66-112 |
| CRP - high sensitivity (high) | 37/507 (7%) | 30/326 (9%) | 0.361 | mg/L | <5 |
| eGFR (low) | 5/508 (1%) | 6/325 (2%) | 0.355 | mL/min/1.73m2 | > 60 |
| Eosinophils (high)  (low) | 14/508 (3%)  0/508 (0%) | 11/324 (3%)  0/324 (0%) | 0.678 | 10^9/L | 0-0.4 |
| ESR (high)  (low) | 40/510 (8%)  0/510 (0%) | 28/323 (9%)  0/323 (0%) | 0.698 | mm/hr | M: 1-20, F and >40 years: 1-23 |
| Gamma GT (high)  (low) | 32/508 (6%)  12/508 (2%) | 18/326 (6%)  4/326 (1%) | 0.765  0.307 | IU/L | F: 6-42, M: 10-71 |
| Globulin (high)  (low) | 2/508 (0%)  14/508 (3%) | 0/326 (0%)  11/326 (3%) | 0.523  0.679 | g/L | 19-35 |
| Haemoglobin (high)  (low) | 6/508 (1%)  13/508 (3%) | 7/324 (2%)  6/324 (2%) | 0.269  0.637 | g/L | F: 115-115, M: 130-170 |
| HCT (high)  (low) | 10/508 (2%)  8/508 (2%) | 12/324 (4%)  1/324 (0%) | 0.182  0.165 |  | F: 0.33-0.45, M: 0.37-0.5 |
| HDL Cholesterol (high)  (low) | 176/508 (35%)  40/508 (8%) | 101/326 (31%)  31/326 (10%) | 0.292  0.446 | mmol/L | F: 1.2-1.7, M: 0.9-1.5 |
| Insulin (high)  (low) | 41/460 (9%)  10/460 (2%) | 27/276 (10%)  7/276 (3%) | 0.695  0.802 | mIU/L | 2.6-24.9 |
| Iron (high)  (low) | 24/508 (5%)  10/508 (2%) | 11/326 (3%)  11/326 (3%) | 0.381  0.258 | μmol/L | F: 6.6-26, M: 10.6-28.3 |
| LDH (high)  (low) | 80/500 (16%)  19/500 (4%) | 70/319 (22%)  10/319 (3%) | **0.034**  0.701 | IU/L | F: 135-214, M: 38-204 |
| LDL Cholesterol (high) | 167/500 (33%) | 116/325 (36%) | 0.5 | mmol/L | <3 |
| Lymphocytes (high)  (low) | 2/508 (0%)  38/508 (7%) | 6/324 (2%)  28/324 (9%) | 0.062  0.599 | 10^9/L | 1.2-3.65 |
| Magnesium (high)  (low) | 2/508 (0%)  1/508 (0%) | 4/326 (1%)  1/326 (0%) | 0.216  >0.999 | mmol/L | 0.6-1.0 |
| MCH (high)  (low) | 4/508 (1%)  8/508 (2%) | 5/324 (2%)  8/324 (2%) | 0.322  0.439 | pg | 26-33.5 |
| MCHC (high)  (low) | 106/508 (21%)  0/508 (0%) | 49/324 (15%)  1/324 (0%) | **0.044**  0.389 | g/L | 300-350 |
| MCV (high)  (low) | 1/508 (0%)  10/508 (2%) | 3/324 (1%)  6/324 (2%) | 0.305  >0.999 | fL | 80-99 |
| Monocytes (high)  (low) | 4/508 (1%)  2/508 (0%) | 2/324 (1%)  0/324 (0%) | >0.999  0.524 | 10^9/L | 0.2-1 |
| MPV (high)  (low) | 8/506 (2%)  0/506 (0%) | 7/324 (2%)  0/324 (0%) | 0.598 | fL | 7-13 |
| Neutrophils (high)  (low) | 8/508 (2%)  30/508 (6%) | 7/324 (2%)  17/324 (5%) | 0.597  0.759 | 10^9/L | 2-7.5 |
| Phosphate (high)  (low) | 13/508 (3%)  53/508 (10%) | 6/326 (2%)  34/326 (10%) | 0.636  >0.999 | mmol/L | 0.87-1.45 |
| Platelet count (high)  (low) | 22/505 (4%)  2/505 (0%) | 15/324 (5%)  2/324 (1%) | 0.864  0.646 | 10^9/L | 150-400 |
| Potassium (high)  (low) | 231/476 (49%)  0/476 (0%) | 91/248 (37%)  0/248 (0%) | **0.003** | mmol/L | 3.5-5.1 |
| RDW (high)  (low) | 12/507 (2%)  26/507 (5%) | 5/324 (2%)  8/324 (2%) | 0.463  0.072 |  | 11.5-15.0 |
| Red cell count (high)  (low) | 14/508 (3%)  17/508 (3%) | 6/324 (2%)  12/324 (4%) | 0.491  0.847 | 10^12/L | F: 3.95-5.15, M: 4.4-5.8 |
| Sodium (high)  (low) | 1/508 (0%)  17/508 (3%) | 2/326 (1%)  8/326 (2%) | 0.564  0.537 | mmol/L | 135-145 |
| Testosterone (high)  (low) | 19/463 (4%)  9/463 (2%) | 7/279 (3%)  7/279 (3%) | 0.306  0.611 | nmol/L | F: 0-1.8, M: 7.6 - 31.4 |
| Thyroid stimulating hormone (high)  (low) | 3/468 (1%)  0/468 (0%) | 4/279 (1%)  1/279 (0%) | 0.434  0.373 | mIU/L | 0.27-4.2 |
| TIBC (high)  (low) | 19/501 (4%)  1/501 (0%) | 5/319 (2%)  5/319 (2%) | 0.088  **0.036** | μmol/L | 41-77 |
| Total protein (high)  (low) | 2/508 (0%)  7/508 (1%) | 0/326 (0%)  9/326 (3%) | 0.523  0.196 | g/L | 63-83 |
| Transferrin saturation (high)  (low) | 9/501 (2%)  79/501 (16%) | 6/319 (2%)  65/319 (20%) | >0.999  0.109 | % | 20-55 |
| Triglycerides (high) | 71/508 (14%) | 44/326 (13%) | 0.918 | mmol/L | <2.3 |
| Troponin I (high) | 4/463 (1%) | 2/279 (1%) | >0.999 | ng/L | < 15.6 |
| Urea (high)  (low) | 1/508 (0%)  1/508 (0%) | 3/326 (1%)  1/326 (0%) | 0.305  >0.999 | mmol/L | 1.7-8.3 |
| Uric acid (high)  (low) | 29/508 (6%)  59/508 (12%) | 19/326 (6%)  27/326 (8%) | >0.999  0.131 | μmol/L | F: 175-363, M: 266-474 |
| White cell count (high)  (low) | 19/508 (4%)  1/508 (0%) | 16/324 (5%)  0/324 (0%) | 0.479  >0.999 | 10^9/L | 3-10 |

**Table S4. Blood investigations by symptom status at baseline and follow-up in individuals with Long Covid.**

|  | **Baseline** | | **p-value** | **Follow-up** | | **p-value** |
| --- | --- | --- | --- | --- | --- | --- |
|  | **Persistent symptoms (n=264)** | **Asymptomatic (n=60)** |  | **Persistent symptoms (n=264)** | **Asymptomatic (n=60)** |  |
| **Bloods** |  |  |  |  |  |  |
| Alanine transferase (high)  (low) | 38/255 (15%)  2/255 (1%) | 11/51 (22%)  1/51 (2%) | 0.294  0.422 | 40/260 (15%)  4/260 (2%) | 5/59 (8%)  0/59 (0%) | 0.215  >0.999 |
| Albumin (high)  (low) | 15/255 (6%)  0/255 (0%) | 3/51 (6%)  0/51 (0%) | >0.999 | 9/260 (3%)  0/260 (0%) | 2/59 (3%)  0/59 (0%) | >0.999 |
| Alkaline Phosphatase (high)  (low) | 7/255 (3%)  7/255 (3%) | 1/51 (2%)  0/51 (0%) | >0.999  0.605 | 9/260 (3%)  5/260 (2%) | 2/59 (3%)  0/59 (0%) | >0.999  0.588 |
| Amylase (high)  (low) | 18/228 (8%)  6/228 (3%) | 4/50 (8%)  0/50 (0%) | >0.999  0.595 | 16/217 (7%)  9/217 (4%) | 5/55 (9%)  1/55 (2%) | 0.777  0.692 |
| Aspartate transferase (high)  (low) | 22/243 (9%)  0/243 (0%) | 4/47 (9%)  0/47 (0%) | >0.999 | 31/248 (12%)  0/248 (0%) | 5/57 (9%)  0/57 (0%) | 0.504 |
| Basophils (high)  (low) | 2/255 (1%)  0/255 (0%) | 0/51 (0%)  0/51 (0%) | >0.999 | 7/258 (3%)  0/258 (0%) | 0/59 (0%)  0/59 (0%) | 0.356 |
| Bicarbonate (high)  (low) | 12/255 (5%)  28/255 (11%) | 5/51 (10%)  4/51 (8%) | 0.175  0.622 | 15/260 (6%)  29/260 (11%) | 6/59 (10%)  4/59 (7%) | 0.243  0.477 |
| Bilirubin (high)  (low) | 5/255 (2%)  0/255 (0%) | 2/51 (4%)  0/51 (0%) | 0.33 | 6/260 (2%)  0/260 (0%) | 6/59 (10%)  0/59 (0%) | **0.012** |
| C-peptide (high)  (low) | 15/228 (7%)  0/228 (0%) | 4/49 (8%)  0/49 (0%) | 0.755 | 19/217 (9%)  1/217 (0%) | 4/55 (7%)  0/55 (0%) | >0.999  >0.999 |
| Calcium (high)  (low) | 3/255 (1%)  4/255 (2%) | 2/51 (4%)  0/51 (0%) | 0.195  >0.999 | 1/260 (0%)  4/260 (2%) | 0/59 (0%)  1/59 (2%) | >0.999  >0.999 |
| Chloride (high)  (low) | 5/255 (2%)  5/255 (2%) | 4/51 (8%)  2/51 (4%) | **0.045**  0.33 | 1/260 (0%)  6/260 (2%) | 0/59 (0%)  0/59 (0%) | >0.999  0.597 |
| Cholesterol (high) | 124/255 (49%) | 24/51 (47%) | 0.879 | 122/260 (47%) | 32/59 (54%) | 0.317 |
| Creatine kinase (high)  (high) | 19/255 (7%)  1/255 (0%) | 6/51 (12%)  0/51 (0%) | 0.276  >0.999 | 33/258 (13%)  2/258 (1%) | 7/58 (12%)  0/58 (0%) | >0.999  >0.999 |
| Creatinine (high)  (low) | 5/255 (2%)  12/255 (5%) | 0/51 (0%)  2/51 (4%) | 0.594  >0.999 | 6/260 (2%)  12/260 (5%) | 0/59 (0%)  3/59 (5%) | 0.597  0.745 |
| CRP - high sensitivity (high) | 25/254 (10%) | 4/51 (8%) | 0.798 | 27/260 (10%) | 3/59 (5%) | 0.321 |
| Eosinophils (high)  (low) | 7/255 (3%)  0/255 (0%) | 0/51 (0%)  0/51 (0%) | 0.605 | 8/258 (3%)  0/258 (0%) | 2/59 (3%)  0/59 (0%) | >0.999 |
| ESR (high)  (low) | 27/255 (11%)  0/255 (0%) | 5/51 (10%)  0/51 (0%) | >0.999 | 23/257 (9%)  0/257 (0%) | 5/59 (8%)  0/59 (0%) | >0.999 |
| Estimated GFR (low) | 4/255 (2%) | 0/51 (0%) | >0.999 | 5/259 (2%) | 1/59 (2%) | >0.999 |
| Gamma GT (high)  (low) | 20/255 (8%)  4/255 (2%) | 1/51 (2%)  1/51 (2%) | 0.22  >0.999 | 17/260 (7%)  4/260 (2%) | 1/59 (2%)  0/59 (0%) | 0.213  >0.999 |
| Globulin (high)  (low) | 0/255 (0%)  7/255 (3%) | 1/51 (2%)  2/51 (4%) | 0.167  0.648 | 0/260 (0%)  7/260 (3%) | 0/59 (0%)  4/59 (7%) | 0.126 |
| Haemoglobin (high)  (low) | 5/255 (2%)  6/255 (2%) | 0/51 (0%)  1/51 (2%) | 0.594  >0.999 | 7/258 (3%)  6/258 (2%) | 0/59 (0%)  0/59 (0%) | 0.356  0.598 |
| HCT (high)  (low) | 8/255 (3%)  4/255 (2%) | 1/51 (2%)  0/51 (0%) | >0.999  >0.999 | 11/258 (4%)  1/258 (0%) | 1/59 (2%)  0/59 (0%) | 0.703  >0.999 |
| HDL Cholesterol (high)  (low) | 82/255 (32%)  27/255 (11%) | 20/51 (39%)  3/51 (6%) | 0.333  0.44 | 76/260 (29%)  28/260 (11%) | 22/59 (37%)  2/59 (3%) | 0.274  0.088 |
| Insulin (high)  (low) | 24/227 (11%)  2/227 (1%) | 6/48 (12%)  0/48 (0%) | 0.62  >0.999 | 21/214 (10%)  6/214 (3%) | 5/55 (9%)  1/55 (2%) | >0.999  >0.999 |
| Iron (high)  (low) | 13/255 (5%)  4/255 (2%) | 1/51 (2%)  1/51 (2%) | 0.479  >0.999 | 6/260 (2%)  7/260 (3%) | 5/59 (8%)  3/59 (5%) | **0.034**  0.401 |
| LDH (high)  (low) | 48/251 (19%)  7/251 (3%) | 10/48 (21%)  1/48 (2%) | 0.842  >0.999 | 58/254 (23%)  8/254 (3%) | 11/58 (19%)  2/58 (3%) | 0.601  >0.999 |
| LDL Cholesterol (high) | 86/248 (35%) | 20/51 (39%) | 0.526 | 91/259 (35%) | 23/59 (39%) | 0.652 |
| Lymphocytes (high)  (low) | 2/255 (1%)  19/255 (7%) | 0/51 (0%)  5/51 (10%) | >0.999  0.57 | 6/258 (2%)  21/258 (8%) | 0/59 (0%)  6/59 (10%) | 0.598  0.608 |
| Magnesium (high)  (low) | 2/255 (1%)  1/255 (0%) | 0/51 (0%)  0/51 (0%) | >0.999  >0.999 | 4/260 (2%)  1/260 (0%) | 0/59 (0%)  0/59 (0%) | >0.999  >0.999 |
| MCH (high)  (low) | 1/255 (0%)  4/255 (2%) | 2/51 (4%)  2/51 (4%) | 0.073  0.263 | 3/258 (1%)  5/258 (2%) | 2/59 (3%)  3/59 (5%) | 0.234  0.171 |
| MCHC (high)  (low) | 52/255 (20%)  0/255 (0%) | 8/51 (16%)  0/51 (0%) | 0.563 | 43/258 (17%)  1/258 (0%) | 5/59 (8%)  0/59 (0%) | 0.157  >0.999 |
| MCV (high)  (low) | 0/255 (0%)  6/255 (2%) | 1/51 (2%)  2/51 (4%) | 0.167  0.624 | 2/258 (1%)  4/258 (2%) | 1/59 (2%)  2/59 (3%) | 0.462  0.31 |
| Monocytes (high)  (low) | 3/255 (1%)  0/255 (0%) | 0/51 (0%)  0/51 (0%) | >0.999 | 2/258 (1%)  0/258 (0%) | 0/59 (0%)  0/59 (0%) | >0.999 |
| MPV (high)  (low) | 6/255 (2%)  0/255 (0%) | 0/51 (0%)  0/51 (0%) | 0.594 | 7/258 (3%)  0/258 (0%) | 0/59 (0%)  0/59 (0%) | 0.356 |
| Neutrophils (high)  (low) | 5/255 (2%)  9/255 (4%) | 1/51 (2%)  3/51 (6%) | >0.999  0.429 | 6/258 (2%)  12/258 (5%) | 0/59 (0%)  5/59 (8%) | 0.598  0.331 |
| Phosphate (high)  (low) | 4/255 (2%)  29/255 (11%) | 2/51 (4%)  8/51 (16%) | 0.263  0.357 | 5/260 (2%)  28/260 (11%) | 1/59 (2%)  5/59 (8%) | >0.999  0.813 |
| Platelet count (high)  (low) | 13/253 (5%)  1/253 (0%) | 3/51 (6%)  0/51 (0%) | 0.738  >0.999 | 13/258 (5%)  0/258 (0%) | 1/59 (2%)  2/59 (3%) | 0.48  **0.034** |
| Potassium (high)  (low) | 114/240 (48%)  0/240 (0%) | 20/42 (48%)  0/42 (0%) | >0.999 | 69/198 (35%)  0/198 (0%) | 20/45 (44%)  0/45 (0%) | 0.235 |
| RDW (high)  (low) | 7/254 (3%) | 3/51 (6%) | 0.38 | 4/258 (2%) | 1/59 (2%) | >0.999 |
|  | 15/254 (6%) | 2/51 (4%) | 0.747 | 8/258 (3%) | 0/59 (0%) | 0.36 |
| Red cell count (high)  (low) | 8/255 (3%)  6/255 (2%) | 1/51 (2%)  2/51 (4%) | >0.999  0.624 | 5/258 (2%)  10/258 (4%) | 1/59 (2%)  2/59 (3%) | >0.999  >0.999 |
| Sodium (high)  (low) | 1/255 (0%)  8/255 (3%) | 0/51 (0%)  2/51 (4%) | >0.999  0.675 | 2/260 (1%)  6/260 (2%) | 0/59 (0%)  2/59 (3%) | >0.999  0.644 |
| Testosterone (high)  (low) | 7/228 (3%)  5/228 (2%) | 3/50 (6%)  0/50 (0%) | 0.393  0.589 | 6/217 (3%)  6/217 (3%) | 1/55 (2%)  1/55 (2%) | >0.999  >0.999 |
| Thyroid stimulating hormone (high)  (low) | 3/230 (1%)  0/230 (0%) | 0/52 (0%)  0/52 (0%) | >0.999 | 3/217 (1%)  0/217 (0%) | 1/55 (2%)  1/55 (2%) | >0.999  0.202 |
| TIBC (high)  (low) | 6/251 (2%)  1/251 (0%) | 1/49 (2%)  0/49 (0%) | >0.999  >0.999 | 4/254 (2%)  4/254 (2%) | 1/58 (2%)  1/58 (2%) | >0.999  >0.999 |
| Total protein (high)  (low) | 1/255 (0%)  5/255 (2%) | 0/51 (0%)  1/51 (2%) | >0.999  >0.999 | 0/260 (0%)  6/260 (2%) | 0/59 (0%)  3/59 (5%) | 0.375 |
| Transferrin saturation (high)  (low) | 4/251 (2%)  46/251 (18%) | 2/49 (4%)  9/49 (18%) | 0.254  >0.999 | 5/254 (2%)  54/254 (21%) | 1/58 (2%)  10/58 (17%) | >0.999  0.59 |
| Triglycerides (high) | 43/255 (17%) | 5/51 (10%) | 0.291 | 37/260 (14%) | 6/59 (10%) | 0.528 |
| Troponin I (high) | 1/228 (0%) | 2/50 (4%) | 0.084 | 0/217 (0%) | 2/55 (4%) | **0.04** |
| Urea (high)  (low) | 0/255 (0%)  0/255 (0%) | 0/51 (0%)  0/51 (0%) |  | 2/260 (1%)  1/260 (0%) | 1/59 (2%)  0/59 (0%) | 0.46  >0.999 |
| Uric acid (high)  (low) | 18/255 (7%)  26/255 (10%) | 4/51 (8%)  5/51 (10%) | 0.771  >0.999 | 16/260 (6%)  21/260 (8%) | 3/59 (5%)  4/59 (7%) | >0.999  >0.999 |
| White cell count (high)  (low) | 11/255 (4%)  0/255 (0%) | 2/51 (4%)  0/51 (0%) | >0.999 | 14/258 (5%)  0/258 (0%) | 1/59 (2%)  0/59 (0%) | 0.32 |

**Table S5: Comparison of MRI metrics between healthy controls and long COVID cohort at baseline.**

| **Metric** | **N**  **total** | **Healthy controls**  **(n=92)** | **Long COVID**  **(n=536)** | **P-value** |
| --- | --- | --- | --- | --- |
| **CARDIAC** | | | | |
| (Field strength independent metrics) | | | | |
| **Global longitudinal strain 3D (%)** | 601 | -14.68 (-15.95, -13.69) | -14.50 (-16.07, -13.08) | 0.475 |
| Missing |  | 13 | 14 |  |
| **RV EF (%)** | 621 | 57.6 (4.5) | 59.1 (4.9) | **0.004** |
| Missing |  | 5 | 2 |  |
| **LV EF (%)** | 625 | 59.5 (56.6, 62.7) | 59.7 (56.8, 62.4) | 0.747 |
| Missing |  | 1 | 2 |  |
| **RV EDV (ml)** (BSA corrected) | 621 | 87 (78 101) | 77 (68 87) | **<0.001** |
| Missing |  | 5 | 2 |  |
| **LV EDV (ml)** (BSA corrected) | 593 | 86 (78, 99) | 80 (70, 90) | **<0.001** |
| Missing |  | 5 | 2 |  |
| (Field strength dependent: 1.5T) | | | | |
| **Global T1 (ms)** | 387 | 968 (962, 988) | 976 (956, 991) | 0.592 |
| Missing |  | 1 | 9 |  |
| (Field strength dependent: 3T) | | | | |
| **Global T1 (ms)** | 224 | 1,179 (1,152, 1,199) | 1,182 (1,159, 1,199) | 0.972 |
| Missing |  | 0 | 7 |  |
| **LIVER** | | | | |
| (Field strength independent metrics) | | | | |
| **cT1 (ms)** | 613 | 709 (667, 748) | 714 (669, 759) | 0.304 |
| Missing |  | 3 | 12 |  |
| **PDFF (%)** | 578 | 1.8 (1.3, 2.6) | 2.6 (1.6, 5.0) | **<0.001** |
| Missing |  | 3 | 1 |  |
| **Liver volume (ml)** | 626 | 1,344 (1,238, 1,550) | 1,420 (1,269, 1,636) | 0.126 |
| Missing |  | 1 | 1 |  |
| **KIDNEY** | | | | |
| (Field strength independent metrics) | | | | |
| **Left volume (ml)** | 624 | 141 (125, 170) | 149 (129, 169) | 0.430 |
| Missing |  | 2 | 2 |  |
| **Right volume (ml)** | 622 | 151 (131, 178) | 149 (132, 168) | 0.392 |
| Missing |  | 2 | 4 |  |
| (Field strength dependent: 1.5T) | | | | |
| **Left cortex T1 (ms)** | 394 | 1,071 (1,033, 1,100) | 1,076 (1,036, 1,127) | 0.186 |
| Missing |  | 1 | 2 |  |
| **Right cortex T1 (ms)** | 395 | 1,058 (1,010, 1,087) | 1,070 (1,025, 1,120) | **0.043** |
| Missing |  | 1 | 1 |  |
| (Field strength dependent: 3T) | | | | |
| **Left cortex T1 (ms)** | 227 | 1,397 (61) | 1,412 (75) | 0.213 |
| Missing |  | 0 | 4 |  |
| **Right cortex T1 (ms)** | 225 | 1,389 (69) | 1,389 (79) | 0.969 |
| Missing |  | 0 | 6 |  |
| **PANCREAS** | | | | |
| (Field strength independent metrics) | | | | |
| **sT1 (ms)** | 590 | 714 (686, 743) | 717 (683, 761) | 0.390 |
| Missing |  | 6 | 32 |  |
| **PDFF (%)** | 607 | 2.11 (1.62, 2.91) | 2.80 (2.10, 4.75) | **<0.001** |
| Missing |  | 4 | 17 |  |
| **SPLEEN** | | | | |
| (Field strength independent metrics) | | | | |
| **Volume (ml)** | 624 | 182 (121, 239) | 182 (146, 240) | 0.248 |
| Missing |  | 1 | 3 |  |
| **LUNG** | | | | |
| (Field strength independent metrics) | | | | |
| **Deep fractional area change (%)** | 582 | 46 (10) | 44 (10) | 0.130 |
| Missing |  | 17 | 29 |  |

**Table S6: Associations between biomarkers and symptom groups in individuals with Long COVID.** Odds ratios (95% CI) and p-values from significant models are reported. The timepoint ‘prediction’ refers to an association of biomarker at baseline and symptom at follow-up. Sample sizes for presenting the outcome (yes/no) refer to observations with no missing data.

| **Symptom group** | **Timepoint** | **MRI metrics (stepwise model)** | **Blood metrics (stepwise model)** | **Combined (multivariable model)** |
| --- | --- | --- | --- | --- |
| **Severe breathlessness** | baseline | Liver fat: OR 1.4 (1.13, 1.74), p=0.002 Pancreatic fat: OR 1.21 (0.98, 1.5), p=0.08 Kidney cortex T1: OR 1.17 (0.95, 1.43), p=0.137 Liver cT1: OR 0.84 (0.67, 1.05), p=0.121 Age: OR: 0.69 (0.56, 0.86), p=0.001 Sex (male): OR 0.59 (0.36, 0.94), p=0.028  Sample size: yes n=161, no, n=281 | TIBC (low): OR 2.04 (0, >2) p=0.979 Transferrin sat (low): OR 1.33 (1.08, 1.63), p=0.007 Cholesterol (high): OR 1.25 (1.01, 1.56), p=0.042 MCHC (high): OR 1.21 (0.98, 1.48), p=0.074 Age: OR 0.79 (0.63, 0.99), p=0.044  Sample size: yes n=150, no, n=248 | Liver fat: OR 1.29 (1.04, 1.58), p=0.018 Transferrin sat (low): OR 1.19 (0.99, 1.44), p=0.067 Cholesterol (high): OR 1.14 (0.93, 1.39), p=0.208 BMI: OR 1.06 (0.86, 1.31), p=0.558 Age: OR 0.75 (0.62, 0.92), p=0.007 Sex (male): OR 0.72 (0.46, 1.13), p=0.15  Sample size: yes n=177, n=307 |
|  | follow-up | Liver volume: OR 1.42 (1.02, 1.99), p=0.037 Kidney volume: OR 0.68 (0.43, 1.05), p=0.082 Age: OR 0.67 (0.48, 0.93), p=0.017  Sample size: yes n=85, n=126 | none |  |
|  | prediction | Liver fat: OR 1.42 (1.08, 1.87), p=0.013 Lungs FAC: OR 1.23 (0.94, 1.6), p=0.126 Age: OR 0.71 (0.53, 0.96), p=0.024 Sex (male): OR 0.47 (0.24, 0.93), p=0.03  Sample size: yes n=79, n=177 | Basophils (high): OR 2.78 (0, >2), p=0.986 Cholesterol (high): OR 1.49 (1.01, 2.22), p=0.046 BMI: OR 1.33 (0.98, 1.79), p=0.065 Age: OR 0.79 (0.58, 1.08), p=0.143 LDH (high): OR 0.76 (0.54, 1.06), p=0.106 LDL Cholesterol (high): OR 0.73 (0.49, 1.08), p=0.113  Sample size: yes n=71, n=156 | Cholesterol (high): OR 1.28 (0.98, 1.67), p=0.075 BMI: OR 1.26 (0.95, 1.68), p=0.105 Liver volume: OR 0.88 (0.66, 1.18), p=0.397 Age: OR 0.79 (0.6, 1.03), p=0.087 Sex (male): OR 0.78 (0.43, 1.43), p=0.42  Sample size: yes n=91, n=197 |
| **Cognitive dysfunction** | baseline | none | Potassium (high): OR 1.36 (1.12, 1.66), p=0.002  Sample size: yes n=218, n=195 |  |
|  | follow-up | Liver cT1: OR 1.48 (1.08, 2.04), p=0.015 Kidney cortex T1: OR 0.75 (0.54, 1.04), p=0.08 Pancreatic fat: OR 0.62 (0.43, 0.9), p=0.011 Sex (male): OR 0.38 (0.18, 0.79), p=0.01  Sample size: yes n=77, no n=120 | none |  |
|  | prediction | none | none |  |
| **Poor HRQoL** | baseline | none | none |  |
|  | follow-up | Liver cT1: OR 1.48 (1.08, 2.04), p=0.015 Kidney cortex T1: OR 0.75 (0.54, 1.04), p=0.08 Pancreas PDFF: OR 0.62 (0.43, 0.9,) p=0.011 Sex (male): OR 0.38 (0.18, 0.79), p=0.01  Sample size: yes n=82, no n=102 | CK (high): OR 0.79 (0.58, 1.08), p=0.14 Sex (male): OR 0.51 (0.26, 1), p=0.05  Sample size: yes n=93, no n=99 | Liver volume: OR 1.12 (0.86, 1.47), p=0.405 Age: OR 1.04 (0.81, 1.32), p=0.771 HDL Cholesterol: OR 1 (0.78, 1.28), p=0.977 Transferrin sat (high): OR 0.93 (0.73, 1.18), p=0.55 BMI: OR 0.91 (0.68, 1.21), p=0.511 Sex (male): OR 0.45 (0.26, 0.8), p=0.006  Sample size: yes n=129, no n=163 |
|  | prediction | none | none |  |

**Figure S1: Example MRI data segmentations used in organ morphology measurements.**

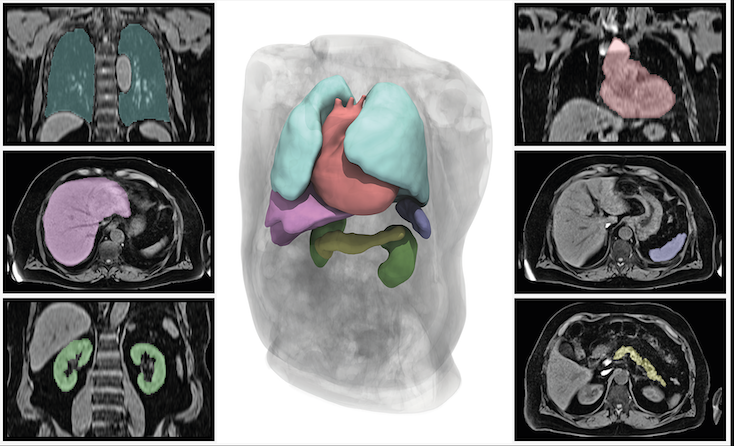

**Figure S2: Self-reported health related quality of life as reported from the EQ-5D-5L instrument showing the UK specific index scores (left) and the visual analogue score (right).**

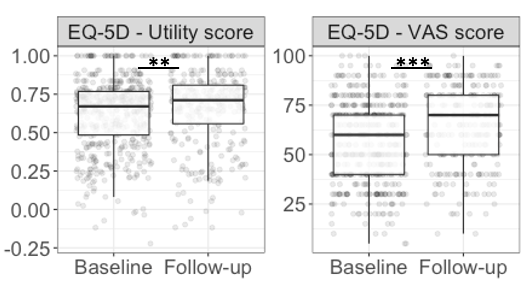

**Figure S3: Dimensions of health from the EQ-5D-5L questionnaire.**

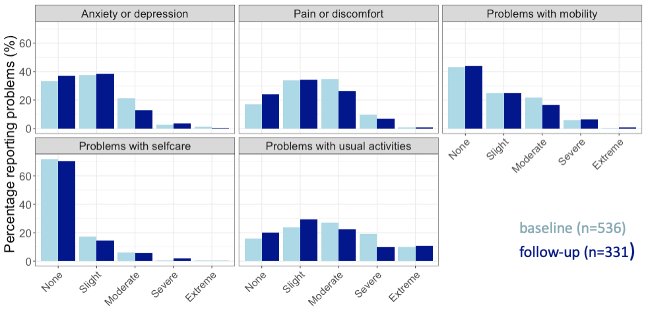

Lighter bars represent baseline scores and darker bars the scores at follow-up. Note that the follow-up group is a subsample of the baseline group.

**Figure S4: Heat maps showing the proportion of those with impairment in individual organs that reported specific symptoms at baseline (left) and follow-up (right).**

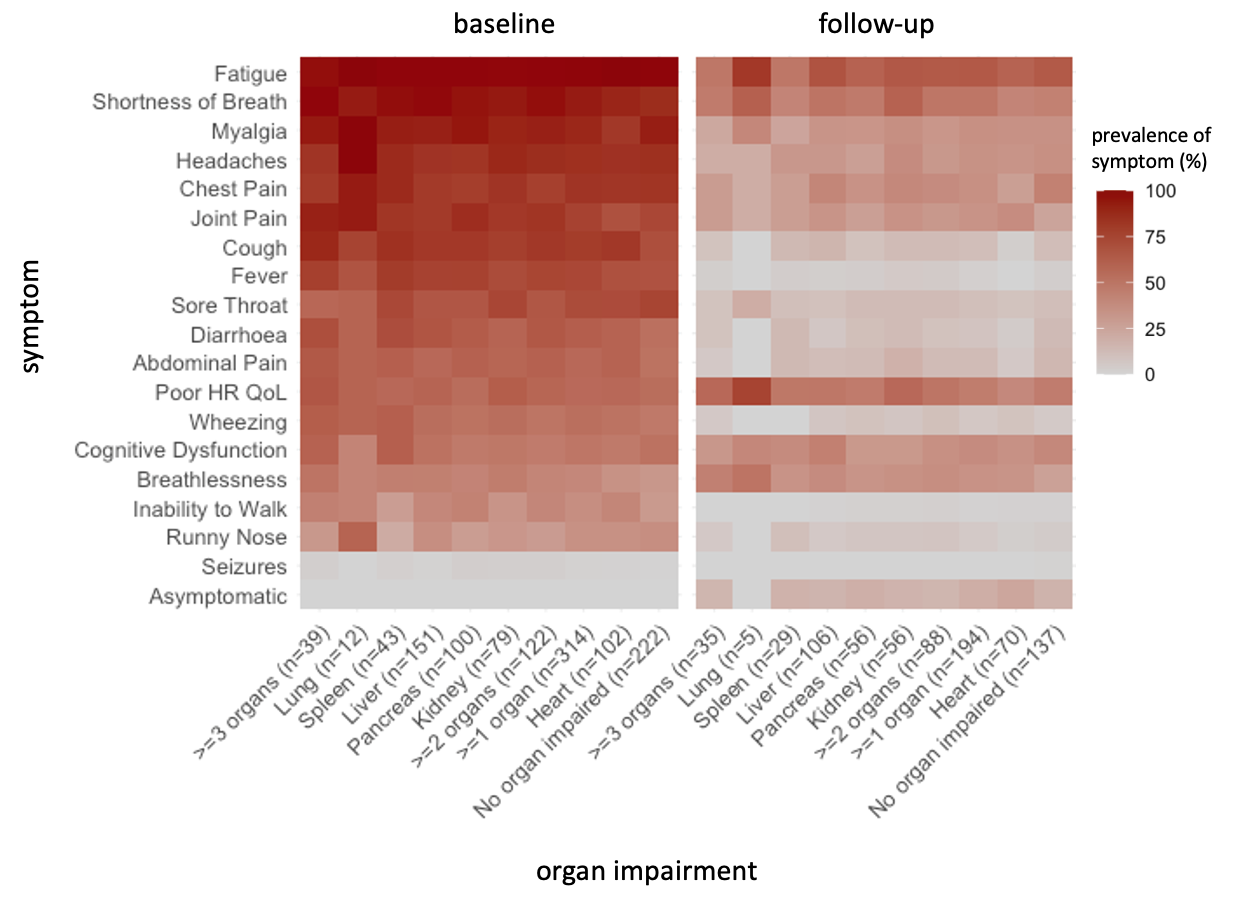

Rows and columns are sorted by mean prevalence of the symptom at baseline. Labels of the x-axis indicate the number of patients presenting with organ impairment at each time point. Shortness of breath is self-reported, and breathlessness is based on Dyspnoea-12. Darker colours indicate a higher proportion as defined by the scale on the right.

**Figure S5: Proportion of cases with liver steatosis by symptom group (systemic, cardiopulmonary, severe breathlessness, brain fog, poor HRQoL).**

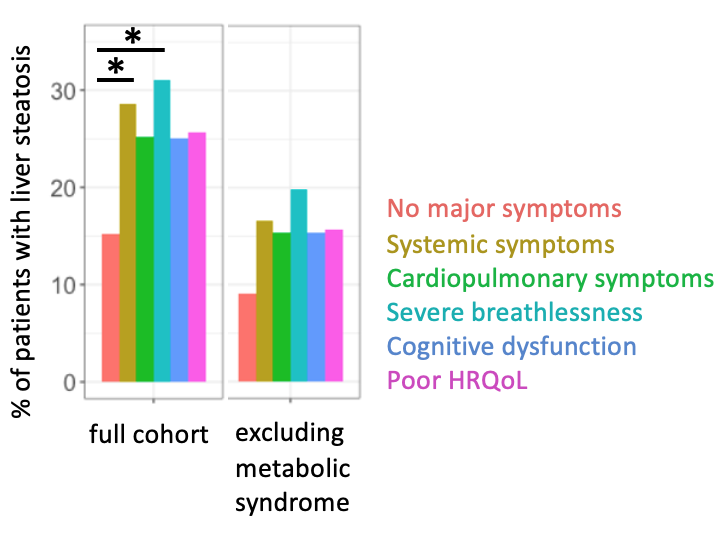

N.B. Prevalence in full cohort at baseline (n=536) and in a subgroup of cases without metabolic syndrome (BMI>=30 or diabetes or hypertension) (n=144).
